# Evaluation of polygenic score for hypertrophic cardiomyopathy in the general population and across clinical settings

**DOI:** 10.1101/2023.03.14.23286621

**Authors:** Sean L Zheng, Sean J Jurgens, Kathryn A McGurk, Xiao Xu, Chris Grace, Pantazis I Theotokis, Rachel J Buchan, Catherine Francis, Antonio de Marvao, Lara Curran, Wenjia Bai, Chee Jian Pua, Tang Hak Chiaw, Paloma Jorda, Marjon A van Slegtenhorst, Judith MA Verhagen, Andrew R Harper, Elizabeth Ormondroyd, Calvin WL Chin, Genomics England Research Consortium, HCM GWAS Collaborators, Antonis Pantazis, John Baksi, Brian P Halliday, Paul Matthews, Yigal M Pinto, Roddy Walsh, Ahmad S Amin, Arthur AM Wilde, Stuart A Cook, Sanjay K Prasad, Paul JR Barton, Declan P O’Regan, RT Lumbers, Anuj Goel, Rafik Tadros, Michelle Michels, Hugh Watkins, Connie R Bezzina, James S Ware

## Abstract

Hypertrophic cardiomyopathy (HCM) is an important cause of morbidity and mortality, with rare pathogenic variants found in about a third of cases (sarcomere-positive). Large-scale genome-wide association studies (GWAS) demonstrate that common genetic variation contributes substantially to HCM risk. Here, we derive polygenic scores (PGS) from HCM GWAS, and multi-trait analysis of GWAS incorporating genetically-correlated traits, and test their performance in the UK Biobank, 100,000 Genomes Project, and across clinical cohorts. Higher PGS substantially increases population risk of HCM, particularly amongst sarcomere-positive carriers where HCM penetrance differs 10-fold between those in the highest and lowest PGS quintiles. In relatives of HCM patients, PGS stratifies risks of developing HCM and adverse outcomes. Finally, PGS strongly predicts risk of adverse outcomes in HCM, with a 4 to 6-fold increase in death between cases in the highest and lowest PGS quintiles. These findings promise broad clinical utility of PGS in the general population, in cases, and in families with HCM, enabling tailored screening and surveillance, and stratification of risk of adverse outcomes.

---

Hypertrophic cardiomyopathy (HCM) is a primary cardiac disease characterized by excessive hypertrophy of the left ventricle with a population prevalence of 0.2%^1^. While many cases follow a benign course, HCM is an important cause of sudden cardiac death in young adults, and progressive disease is complicated by arrhythmia, stroke, and heart failure^2,3^. Although HCM has classically been considered a Mendelian disease, a causal rare variant is identified in only a third of cases^4,5^, with population studies highlighting the incomplete penetrance and variable expressivity of such variants^6,7^. Recent genome-wide association studies (GWAS) demonstrated that common variants contribute substantially to HCM risk (SNP *h*^*2*^ 0.29), identified many contributory loci, and highlighted the complex genetic architecture of HCM^8-10^. Polygenic scores (PGS) summarize the cumulative risk arising from common variants and may provide important utility for population risk prediction and prognostication^9,11^. Still, it remains unclear whether PGS can inform risk of HCM and clinical outcomes across broad clinical and population settings. The aim of this study is to develop and evaluate a PGS for HCM, assessing utility for stratification of both disease risk and severity in (1) individuals diagnosed with HCM, (2) relatives of affected individuals who would currently be recommended to undergo screening and long-term surveillance, and (3) the general population, including individuals carrying disease-associated rare variants such as those that might be identified as secondary findings.

## Results

### Generation and evaluation of a polygenic risk score for hypertrophic cardiomyopathy in the general population

PGS was generated using the largest published GWAS comprising a total of 5,900 unrelated HCM cases and 68,359 controls of European ancestry from 7 cohorts (PGS_GWAS_), and multi-trait analysis of GWAS (MTAG) incorporating the HCM GWAS with GWAS of 3 genetically-correlated cardiac magnetic resonance imaging (CMR) traits (left ventricular [LV] concentricity, LV end-systolic volume [LVESV], and LV circumferential strain) in 36,083 European ancestry participants in the UK Biobank (UKB) (PGS_MTAG_) (Figure 1)^10^. In an independent cohort of 343,182 unrelated White British ancestry participants in UKB, PGS_MTAG_ was associated with risk of HCM (OR per PGS_MTAG_ SD 2.34 [95% CI 2.12 to 2.59,

**Figure 1:**
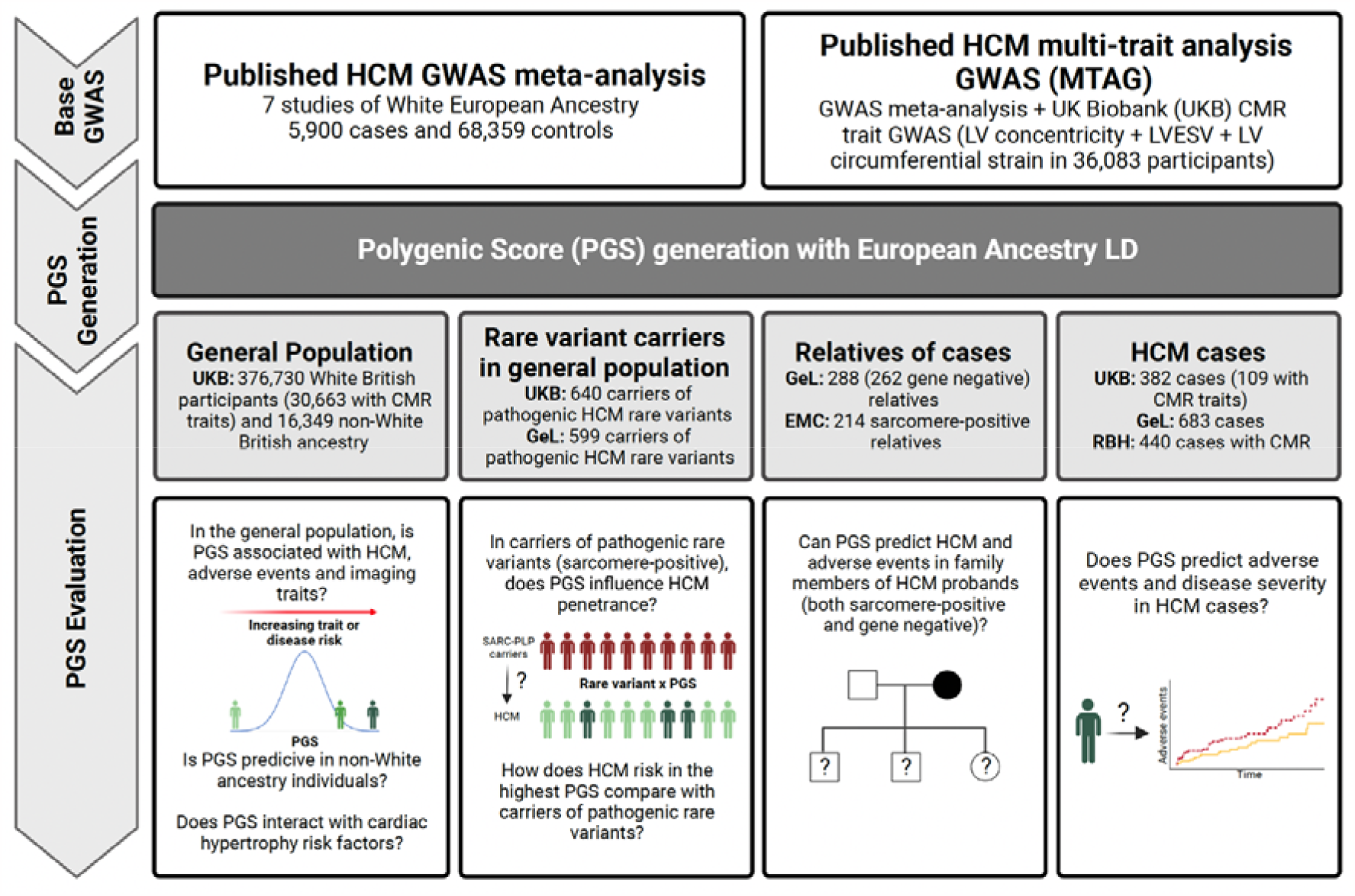
Study overview. Bayesian genome-wide PGS were generated from a published European-ancestry hypertrophic cardiomyopathy (HCM) GWAS meta-analysis of seven case-control studies (comprising 5,900 cases and 68,359 controls; PGS_GWAS_), and multi-trait analysis of GWAS (analyzing HCM with three genetically-correlated quantitative traits measured using cardiac MRI [CMR] in 36,083 UKB participants: LV concentricity, LV end systolic volume and LV circumferential strain; PGS_MTAG_)^10^. The value of PGS to support clinical decision making was evaluated across three key settings: in the general population (including among carriers of pathogenic rare variants in HCM-causing genes [sarcomere-positive] that might be returned as secondary findings), in relatives of HCM probands currently recommended to undergo cascade screening and surveillance, and in confirmed HCM cases under longitudinal follow up. UKB – UK Biobank, GeL – 100,000 Genomes Project; EMC – Erasmus Medical Centre, Netherlands; RBH – Royal Brompton Hospital, UK. LV – left ventricle/ventricular, LVESV – LV end-systolic volume.

P<2×10^−16^]), provided better predictive performance than PGS_GWAS_ (Table S1), and was therefore used for all subsequent analyses unless otherwise stated (Figure S1). The distribution of PGS in the UKB population-based cohort is shown in Figure 2A. Among HCM cases, 75.1% (95% CI 71.4 to 79.5%) have a PGS above the population mean, while those with a PGS greater than one standard deviation above the mean accounted for 46.4% (95% CI 41.2 to 51.7%) of cases (Figure 2B). Phenome-wide association study (pheWAS) of 1,839 clinical diagnoses in the UKB identified PGS associations with hypertension and metabolic phenotypes (dyslipidemia, and type 2 diabetes) and an inverse association with heart failure^9^(Figure 2C). Mendelian randomization highlighted causal influence of blood pressure and body mass index as previously demonstrated^8^, and no significant associations with lipid and glycaemic traits (Table S2 and S3).

**Figure 2:**
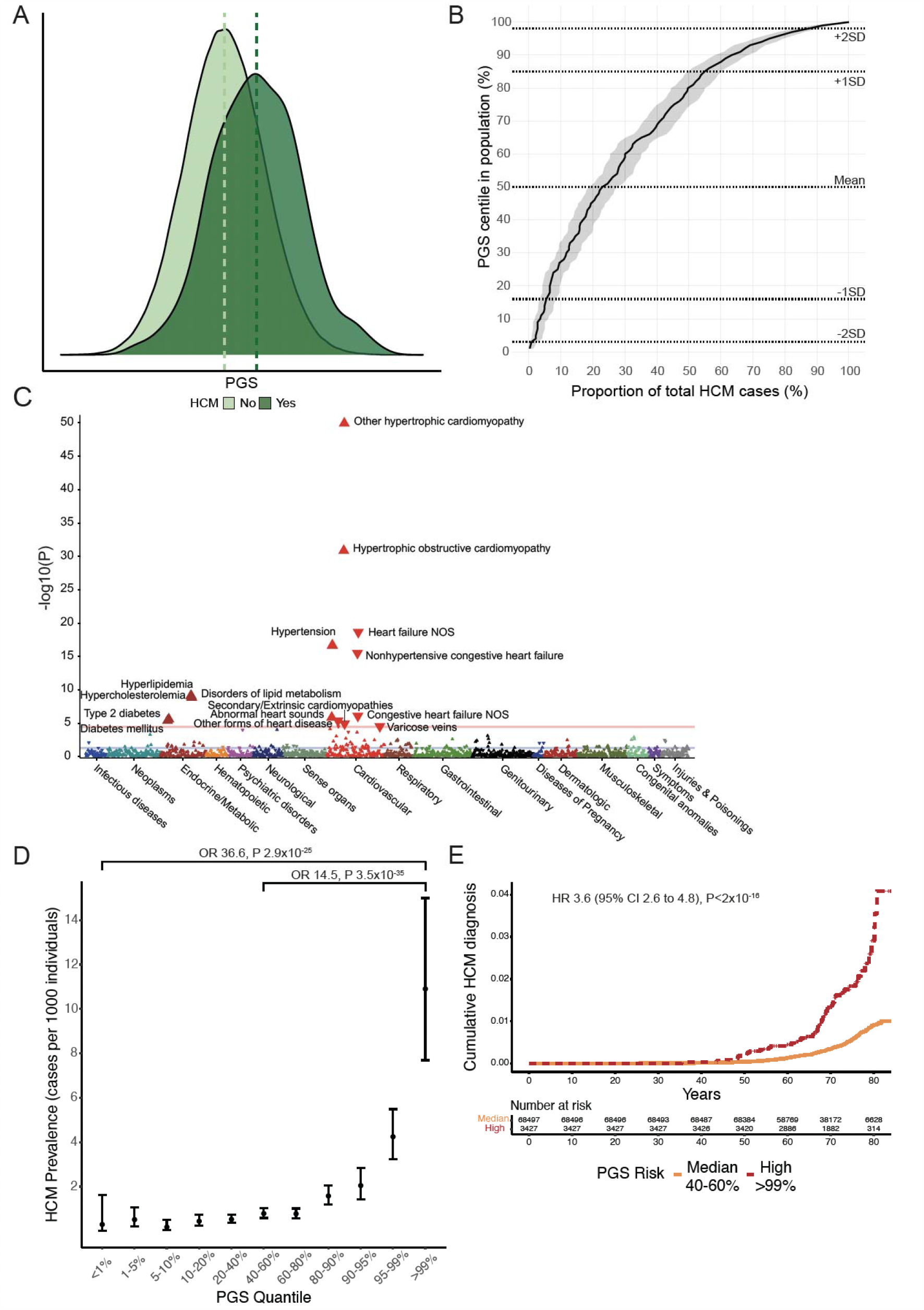
HCM PGS is associated with HCM disease status in the UK Biobank. To validate the PGS, we analysed associations with PGS in the UKB population. (A) PGS_MTAG_ distribution in 374,845 UKB participants with and without HCM demonstrating higher PGS in those with HCM. (B) Cumulative curve of HCM cases ranked across PGS centiles. For example, approximately 75% of HCM cases have a PGS above the 50^th^ centile. Dashed lines represent median, ±1 PGS SD and ±2 PGS SD. (C) Manhattan plot of HCM PGS phenome-wide association study in UKB, showing associations with cardiovascular and metabolic phenotypes. ICD-9 and ICD-10 diagnostic codes are mapped to Phecode Map version 1.2. Mapped phenotypes exceeding phenome-wide significance threshold (P 2.7×10^−5^, red line) are labelled. Blue line indicates nominal significance (P<0.05). Direction of triangle indicates the direction of effect of the PGS association. (D) HCM prevalence and risk in UKB across the spectrum of PGS, demonstrating substantially higher HCM prevalence in individuals with the highest PGS. (E) Cumulative hazards for lifetime diagnosis of HCM stratified by high (highest centile – red) and median (middle quintile – orange) PGS risk in UKB.

Having demonstrated associations between PGS and HCM risk, we evaluated effect sizes and considered clinical utility for disease prediction. In the general population, individuals with PGS in the highest centile (prevalence 10.9 cases/1000 individuals) had substantially higher risks of HCM compared with those in the median (prevalence 0.8 cases/1000, OR 14.5 [95% CI 9.5 to 22.2], P 3.5×10^−35^) and lowest centiles (prevalence 0.3 cases/1000, OR 36.6 [95% CI 18.6 to 72.2], P 2.9×10^−25^) (Figure 2D and 2E, Figure S2, Table S4).

Exploring the role of polygenic risk on expressivity of an HCM phenotype in 30,663 White British ancestry UKB participants who underwent CMR, PGS_GWAS_ was associated with traits that are classically seen in HCM^6^: increased cardiac hypertrophy (maximum LV wall thickness [maxLVWT]: +0.13mm per PGS SD, P 1.1×10^−80^; highest vs. lowest PGS centile: 9.8 vs. 9.1mm, P 9×10^−9^), increased cardiac contractility (LV ejection fraction [LVEF]: +0.6%, P 2.7×10^−64^; 61.3 vs. 57.7%, P 2.7×10^−13^), reduced chamber volumes (LV end-diastolic volume [LVEDV] [-2.0ml, P 1.2×10^−46^; 142.1 vs. 154.9ml, P 8.3×10^−8^]; and LVESV [-1.7ml, P 6.9×10^−80^; 55.9 vs. 66.4, P 2.7×10^−10^]), all of which were biventricular in nature (Figure S3, Tables S5 and S6), and persisted when excluding participants with HCM (Table S7).

### Evaluating and improving HCM PGS performance in non-White ancestry populations

PGS derived from one ancestry underperform when applied to different or more diverse ancestral populations^12-15^. We adapted the European PGS by applying LD ancestry-specific references^16^, and evaluated its performance for HCM status in 16,349 UKB participants of non-White British ancestry (7,542 South Asian, 7,348 African and 1,459 Chinese ancestry), and for CMR quantitative traits in a subset. PGS distributions differed in the different ancestry groups (ANOVA P <2×10^−16^, Tukey adjusted P <2.5×10^−8^ for between group comparisons), with PGS highest in African ancestry (HCM prevalence 0.4%), and lowest in South Asian ancestry (HCM prevalence 0.1%) (Figure S4). While PGS captured differences between ancestries, within each ancestral group PGS performance was poorer (South Asian [9 HCM cases, OR per PGS SD 1.82, P 0.068]; African [27 cases, OR per PGS SD 1.21, P 0.35]; insufficient Chinese ancestry cases to allow estimation [2 cases]) than in White British (OR per PGS SD 2.34, P<2×10^−16^).

To improve cross-ancestral polygenic prediction we performed GWAS in an unrelated Chinese ancestry cohort of 174 HCM cases and 776 controls recruited from Singapore (no individual SNPs reaching genome-wide significance, Figure S5), and combined them with the existing European ancestry HCM GWAS to generate a cross-population PGS^16^ (PGS_Chinese_). PGS_Chinese_ was associated with LV volumetric (LVEDV: change per PGS SD -3.6ml, P 0.028) and wall thickness traits (maxLVWT: +0.28mm, P 0.017; above vs. below mean: 9.5 vs. 8.6mm, P 0.008), which were not present when using the European only PGS (Figure S4).

### PGS modulates the penetrance of HCM-causing rare variants

Amongst 318,945 UKB participants with whole-exome sequencing (WES), there were 640 carriers of pathogenic or likely pathogenic variants in 8 genes encoding components of the cardiac sarcomere (*MYBPC3, MYH7, TNNT2, TNNI3, TPM1, ACTC1, MYL3*, and *MYL2*) (“sarcomere-positive”), and a total of 336 HCM cases (43 sarcomere-positive: penetrance 6.7% [95% CI 4.9 to 8.9%]; 293 sarcomere-negative: prevalence 0.09% [95% CI 0.008 to 0.001%]). Among the UKB population, there was no difference in mean PGS in sarcomere-positive and -negative participants (P 0.60) (Figure S6) arguing against any unmeasured relationship between rare sarcomeric variants and common variant polygenicity that might arise from selective ascertainment. PGS was associated with HCM in both sarcomere-positive (OR per PGS SD 2.35, P 1.1×10^−6^) and sarcomere-negative participants (OR per PGS SD 2.15, P<2×10^−16^).

To investigate the effect of PGS on penetrance of rare HCM-causing pathogenic variants, we further evaluated the PGS in the 640 unrelated sarcomere-positive individuals in the UKB (Figure 3A). Penetrance by middle-older age (median age 72 [IQR ±13 years]) in sarcomere-positive individuals was markedly greater in the highest quintile (HCM penetrance 17.2% [95% CI 10.8 to 25.3%]) when compared with the median (5.7% [95% CI 2.1 to 12.0%]; highest vs. median quintile OR 3.69 [95% CI 1.46 to 10.67], P 0.009) and lowest quintiles (2.3% [95% CI 0.5 to 6.6%]; highest vs. lowest quintile OR 9.56 [95% CI 2.95 to 43.89], P 7.3×10^−4^) (Figure 3B). In time-to-event analyses, the risk of HCM diagnosis (HR 6.54 [95% CI 2.6 to 16.5], P 6.6×10^−5^) and adverse HCM outcomes (HR 1.56 [95% CI 1.1 to 2.34], P 0.029) were similarly greater in the highest compared with median quintile (Figure 3C, Figure S6).

**Figure 3:**
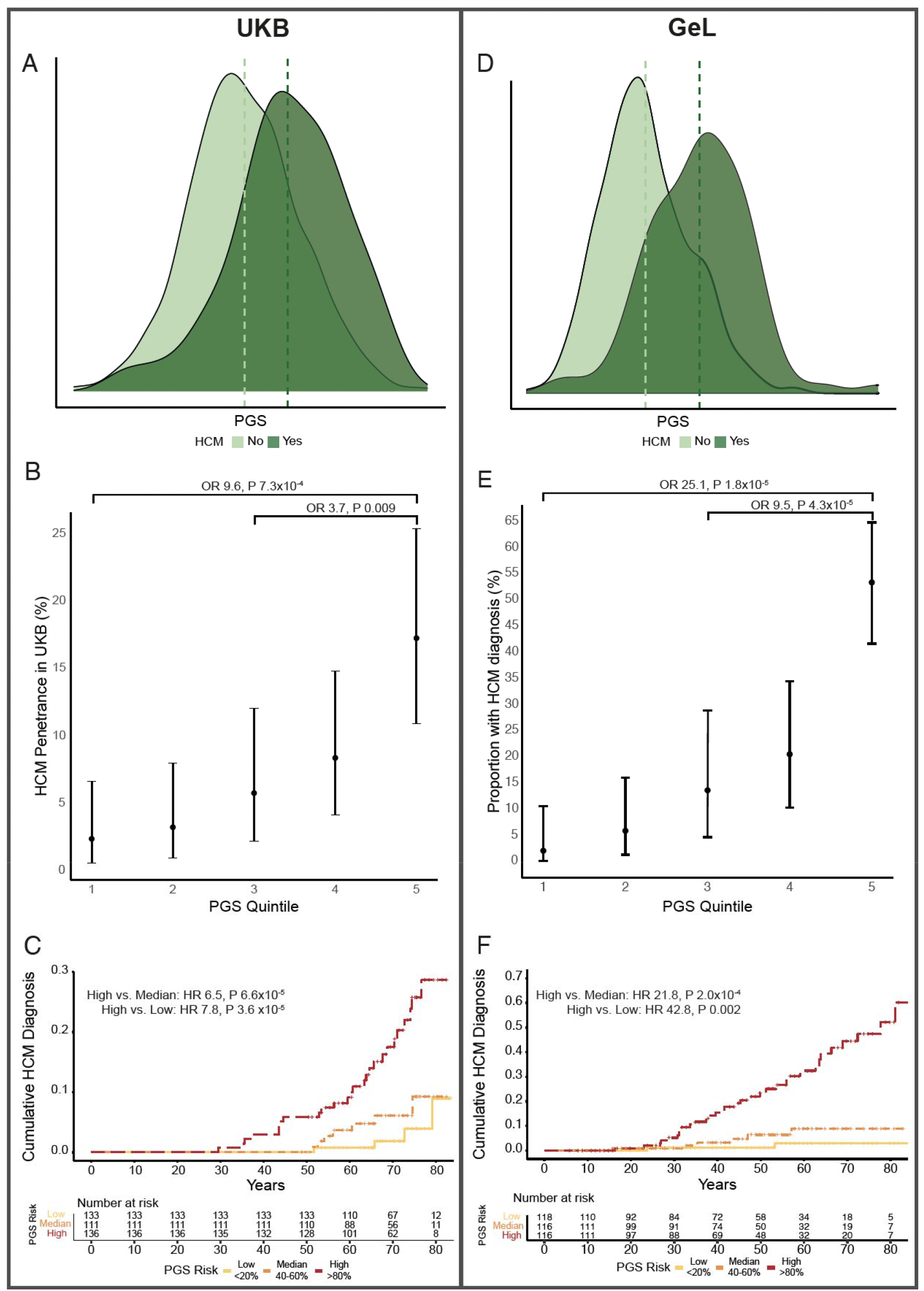
PGS modulates HCM penetrance in carriers of rare pathogenic variants in HCM-associated genes. UKB (A-C) represents a broadly unselected population, as participants were not recruited based on genotype of phenotype. GeL (D-F) comprises a mix of participants recruited based on cardiomyopathy, and participants recruited with other rare diseases, cancer, or as relatives of patients with rare disease. (A & D) show the PGS distribution in 640 sarcomere-positive UKB participants and 599 GeL participants with and without HCM, validating that PGS is higher in cases than controls. B (UKB) and E (GeL) show the proportion of sarcomere-positive individuals that are diagnosed with HCM, stratified by PGS quintile. C (UKB) and F (GeL) show time to HCM diagnosis in highest, median and lowest quintiles, showing that those with higher PGS are at increased risk of HCM, and develop disease earlier, which is important for lifetime burden of disease morbidities.

We confirmed the modulatory role of PGS on rare variants in 100,000 Genomes Project (GeL)^17^ (Figure 3D), a study that recruited individuals with rare diseases (including cardiomyopathies) and their relatives. Since a small proportion of the GeL cohort were ascertained based on cardiomyopathy, we cannot use this data to directly quantify penetrance, but can nonetheless assess the effect size for PGS in combination with a rare variant on disease risk in this cohort. There were 599 sarcomere-positive participants, with 72 HCM cases (proportion affected 12.0% [95% CI 9.6 to 15.0%]). PGS_GeL_ (generated from MTAG summary statistics leaving out the GeL cohort) was associated with prevalent HCM (OR per PGS_GeL_ SD 3.53 [95% CI 2.59 to 4.80], P 9.8×10^−16^, OR per PGS_GeL_ decile 1.60 [95% CI 1.41 to 1.85]). Sarcomere-positive individuals with high PGS_GeL_ (highest quintile) were more than 9 times as likely to have been ascertained as cases compared with the median (OR 9.50 [95% CI 3.60 to 32.6], P 4.3×10^−5^), and 25 times as likely compared with the lowest (OR 25.1 [95% CI 7.3 to 160.3], P 1.8×10^−5^) quintile (Figure 3E). The hazard of HCM diagnosis was higher in the highest quintile than at the median (HR 21.8 [95% CI 4.8 to 98.6], P 0.0002) and lowest (HR 42.8 [95% CI 5.0 to 364.0], P 0.002) quintiles (Figure 3F). Finally, of 527 sarcomere-positive individuals without a diagnosis of HCM on recruitment to GeL, a total of 7 were diagnosed with HCM on follow-up, 5 of whom had PGS_GeL_ in the highest quintile.

### Comparing HCM risk in participants with high PGS risk vs. pathogenic rare-variant carriers

It has been suggested for several diseases that extreme PGS risk confers a similar magnitude of increased risk as the presence of Mendelian pathogenic variants (for example, familial hypercholesterolemia for coronary artery disease, and *BRCA1*/*2* for breast cancer^12,18,19^). In the UKB, HCM risk in those at the uppermost PGS extreme, defined as the top 0.25% (1 in 400 – a frequency comparable to population estimates of pathogenic HCM rare variants^6^) was substantially greater compared with the median PGS (OR 18.1 [95% CI 10.0 to 32.9], P 1.6×10^−21^), though the risk of HCM and severity of imaging traits remained greater in sarcomere-positive individuals compared with those with extreme PGS risk alone (OR 5.4 [95% CI 2.8 to 11.6], P 2.2×10^−8^) (Table S8). These findings suggest that while genetic HCM risk is highest among carriers of rare pathogenic variants, PGS accounts for an important component of risk in sarcomere-negative individuals.

### PGS predicts risk of HCM in the relatives of HCM probands

Understanding the penetrance of HCM in relatives of probands will have important implications on clinical practice (e.g. screening, and longitudinal surveillance). We sought to assess whether PGS modulates penetrance in relatives of sarcomere-positive HCM cases, and stratifies risk in gene negative families, in two cohorts.

GeL was initially designed to evaluate genetically-unexplained rare disease through the recruitment of cases and their relatives, and therefore the cohort has a higher proportion of genetically unexplained sarcomere-negative cases than in the clinical setting (pathogenic rare variants were identified in 94 of 919 HCM cases). In all, 288 relatives of 193 HCM index cases (26 relatives of 14 gene-positive HCM cases) were recruited, of whom 116 had prevalent HCM and 6 were diagnosed with HCM during follow-up. PGS_GeL_ was higher in probands (P<2×10^−16^) and affected relatives (P 3.6×10^−6^) compared with unaffected relatives, with no difference between probands and affected relatives (P=0.99). PGS was associated with increased risk of HCM (HCM OR per PGS_GeL_ SD 1.74 [95% CI 1.37 to 2.21] P 5.1×10^−6^; Highest vs. Median quintile: OR 4.17 [95% CI 1.78 to 10.5], P 0.0015) (Figure 4A). Of 178 relatives who did not have a diagnosis of HCM on recruitment, 6 were diagnosed on follow-up (mean 5.1 years), all with PGS in the highest quintile.

**Figure 4:**
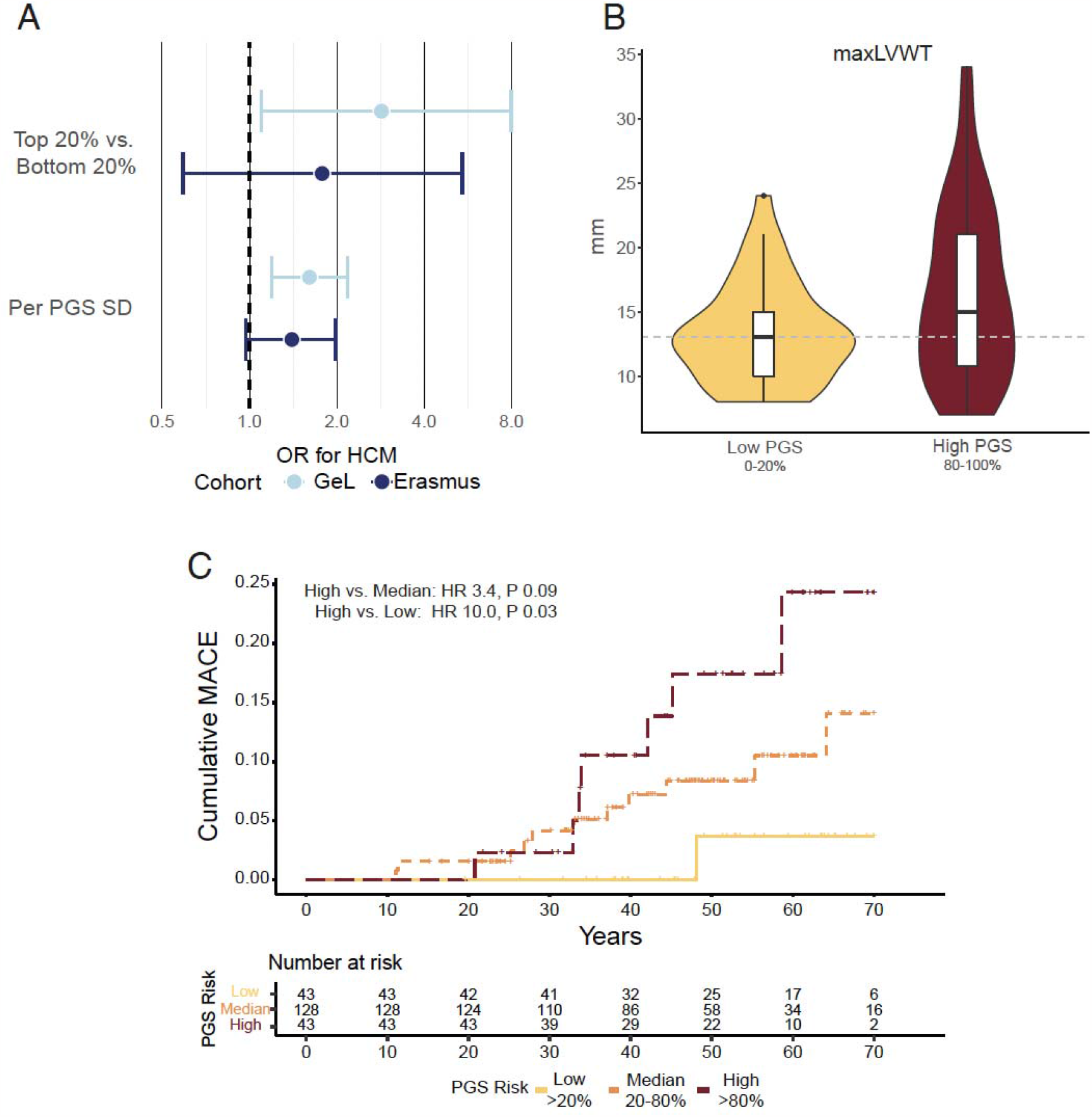
PGS associates with HCM risk and adverse outcomes in relatives of HCM cases. To evaluate applications of PGS in families undergoing screening and surveillance for HCM, we studied relatives of HCM cases in two cohorts, 100,000 Genomes Project [GeL] and Erasmus Medical Centre [EMC] cohort. The GeL cohort was predominantly SARC-Neg (281 of 288) while the entire EMC cohort was sarcomere-positive (214 relatives). (A) OR for HCM among relatives of HCM probands in the two cohorts, stratified by PGS_GeL_. (B) Violin and box and whisker plot of maximum LV wall thickness (maxLVWT) in sarcomere-positive relatives stratified by highest (N=40) and lowest (N=38) PGS_EMC_ quintiles. Dashed line indicates 13mm cut-off used for guideline diagnosis of HCM in relatives of individuals with HCM. (C) Cumulative major adverse cardiovascular events (MACE) among 214 sarcomere positive relatives of HCM index patients stratified by PGS_EMC_ above or below the median. MACE was defined as a composite of septal reduction therapy, cardiac transplantation, aborted cardiac arrest, appropriate defibrillator shock, or sudden cardiac death. To avoid inflation of PGS performance resulting from sample overlap, PGS were re-derived from GWAS leaving out the cohort that the PGS was being evaluated in (GeL – PGS_GeL_, EMC – PGS_EMC_).

The Erasmus Medical Centre (EMC) cohort comprises 214 relatives of 184 index HCM cases, all carriers of rare pathogenic variants in sarcomere-encoding genes. After clinical evaluation, 135 relatives were found to have HCM. Although the PGS_EMC_ (derived using HCM MTAG omitting EMC cohort) was not significantly associated with HCM in relatives (OR 1.36 per PGS_EMC_ SD [95% CI 0.96 to 1.91], P 0.081) (Figure 4A), it was associated with increased maxLVWT (+1.4mm per PGS_EMC_ SD [95% CI 0.6 to 2.1mm], P 5.0×10^−4^; highest vs. lowest quintile: +3.5mm [95% CI 1.26 to 6.41], P 0.0035) and, importantly, with increased risk of MACE after study enrollment (HR per PGS_EMC_ SD 1.74 [95% CI 1.03 to 2.91], P 0.036; highest vs. lowest quintile HR 17.7 [95% CI 0.9 to 347], P 0.058), prevention of which is the primary motivation for cascade screening and early diagnosis (Figure 4B and 4C, Figure S7).

### PGS as a risk predictor in individuals with HCM

Although many individuals with HCM have a relatively benign disease course, several clinical risk factors are associated with adverse outcomes including cardiovascular death, and risk stratification, especially for preventable sudden death, remains an urgent clinical need. We sought to investigate whether PGS was associated with adverse outcomes and clinical features of severity in individuals with HCM. In 382 HCM cases in the UKB, a PGS in the highest quintile was associated with increased risk of death and adverse cardiovascular outcomes after HCM diagnosis (death: HR highest vs. lowest quintile 3.88 [95% CI 1.33 to 11.29], P 0.013; adverse outomces (HCM composite): HR 3.50 [95% CI 1.74 to 7.03], P 4.4×10^−4^) (Figures 5A and 5B, Figure S8). In 683 HCM cases in GeL, cases in the highest quintile had a 6-fold increased risk of death after HCM diagnosis (HR 6.30 [95% CI 2.68 to 14.78], P 1.4×10^−6^) (Figure 5C, Figure S8). In 101 sarcomere-positive HCM cases from a clinical cohort (), higher PGS was associated with a more severe hypertrophic phenotype (maxLVWT: per PGS SD +1.6mm [95% CI 0.61 to 2.63], P 0.002; LV mass: per PGS SD +13.8g [95% CI 1.5 to 26.2], P 0.03) (Figure S8, Table S9).

**Figure 5:**
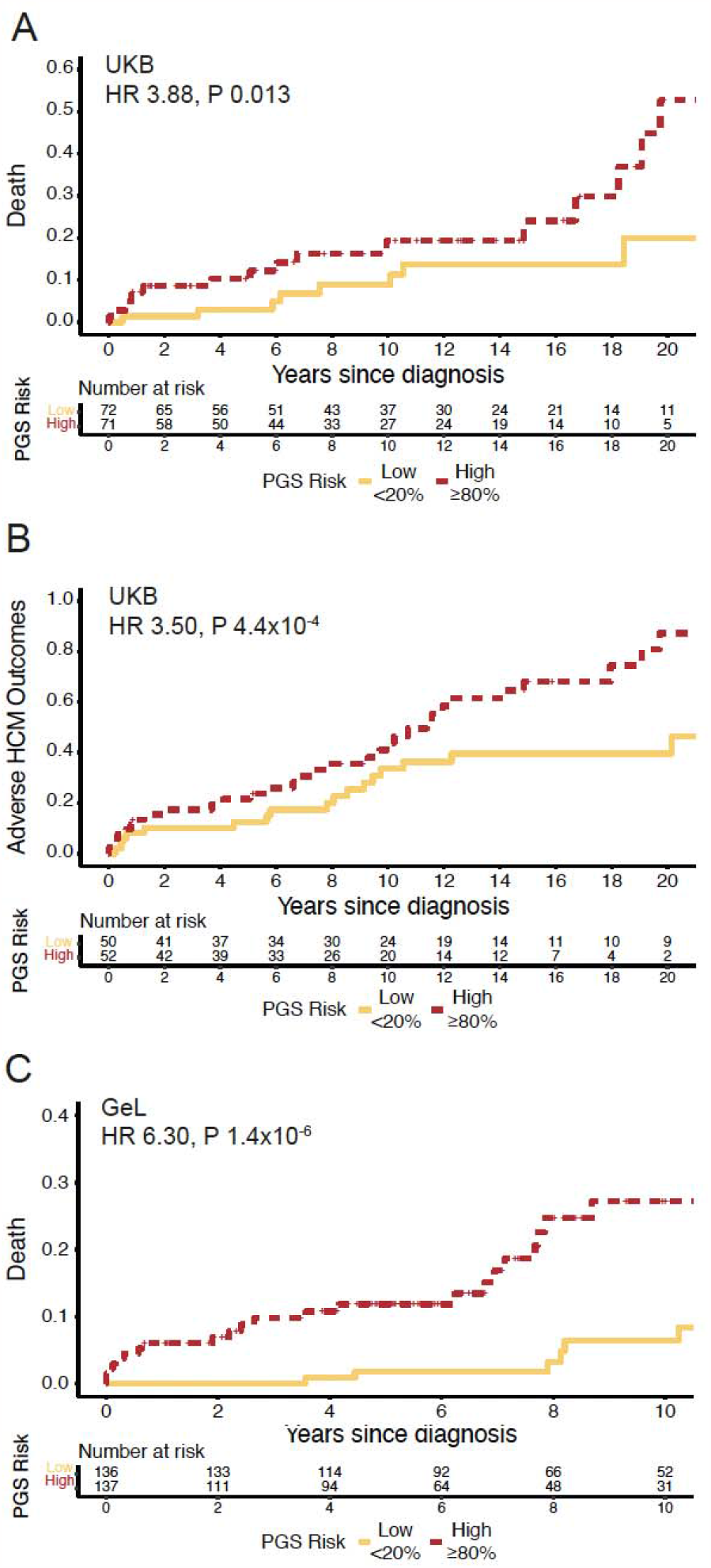
PGS stratifies risk of death and adverse outcomes in individuals with HCM. Cumulative all-cause mortality and adverse HCM outcomes after HCM diagnosis in 382 HCM cases from the UKB (A and B) and cumulative all-cause mortality in 683 HCM cases from GeL (C), stratified by PGS in the highest and lowest quintiles. Adverse HCM outcomes includes death, stroke, cardiac arrest, ICD implantation, septal reduction therapy (alcohol-septal ablation or surgical myectomy), LVAD implantation or cardiac transplantation.

## Discussion

In this study, we generate a PGS for HCM and validate it across several independent populations, showing associations with categorical disease status, quantitative traits that define HCM and describe disease severity, and, most importantly, adverse cardiovascular events. We demonstrate broad potential clinical utility for PGS across a range of settings. Notably, PGS robustly stratifies penetrance in carriers of rare pathogenic variants in sarcomere-encoding genes and identifies those in the general population at highest risk of developing HCM, associates with HCM risk in relatives of HCM probands, and acts as a novel risk marker for survival and adverse events in individuals with HCM.

Findings from this study emphasize the importance of the polygenic contribution to HCM disease risk, classically considered a Mendelian disease caused by rare variants in sarcomere-encoding genes^4,20^. Among individuals with HCM, the recognition of polygenic, rather than sarcomeric HCM, will be of diagnostic importance, with potential implications on clinical management, reproductive counselling, and family screening^21^

One of the key challenges in clinical practice remains understanding the variable penetrance and expressivity that characterize rare variants in HCM-causing sarcomeric genes^2,3^. In relatives of patients with HCM who have inherited a pathogenic variant, clinical screening and life-long surveillance from childhood is recommended, though many will not manifest until later life, if at all, and many of those who do manifest will follow a benign course without major adverse events. Increasingly, pathogenic variants are being identified in individuals with no personal or family history of HCM, as secondary findings through opportunistic screening alongside genetic testing for other indications^22^. We show that PGS has large and clinically meaningful effects (estimated to be approximately 10-fold when comparing quintiles but larger still at more extremes of distribution) in carriers of rare HCM-causing variants, that we expect would translate to effective risk-adjusted strategies for HCM screening and surveillance. Furthermore, while current medical treatments are only indicated in individuals with established HCM, any future development of therapies in the prevention of cardiac hypertrophy in at risk or genetically susceptible individuals could be of particular significance in the groups with highest risk of disease penetrance. This is especially true given that both rare variants and PGS are measurable before clinical phenotypes of HCM develop.

In relatives of individuals with HCM, PGS risk modulates HCM penetrance in carriers of rare variants and stratifies HCM risk in gene negative cases. Relatives with higher PGS were more likely to have HCM, adverse outcomes, and increased wall thickness, with similar magnitudes of risk in relatives of both gene positive and negative HCM. Directly quantifying PGS with genotyping arrays may guide ongoing surveillance strategies in close relatives of affected individuals, though further prospective work is required.

Within the general population, although PGS confers a substantially increased risk of HCM at the extremes of distribution (OR 15 for the highest centile compared with median), this risk was considerably lower than the risk arising from pathogenic rare variants (OR 79).

However, PGS risk increases on a continuum and potentially affects a larger proportion of the population than rare sarcomeric variants (for example, thresholding the highest centile). Our population estimates of PGS performance in the UKB are limited by recruitment targeting participants of middle age, with survival bias and incomplete ascertainment of cases likely to result in underestimation of the true effect of PGS in the general population. The applicability of routine and widespread use of PGS for disease screening remains uncertain^23^. As with any population screening approach, targeting PGS screening to individuals already at higher risk based on non-genetic factors can have a large impact on the numbers needed to test.

Among individuals with HCM, disease expressivity and prognosis is highly variable^4,20,24^. We demonstrate that PGS can stratify risk of serious adverse events in individuals with HCM, including a roughly 4 to 6 -fold difference in risk of death when comparing those with PGS in the highest and lowest quintiles. Despite the use of current clinical risk predictors of adverse outcomes^3^many individuals do not benefit from interventions aimed at reducing this (e.g. ICD implantation^25^). The addition of PGS to existing clinical risk factors will be an important area for future research.

One of the main limitations of this and other PGS is that it has been derived from and extensively tested in European ancestry populations only. Despite this, we show that PGS stratifies ancestral risk, with the highest PGS found in African ancestry groups where the prevalence of unexplained LV hypertrophy is known to be highest^26-29^. Within each non-European ancestry group in the UKB performance is reduced compared to European ancestry performance, although it still associates with HCM-related cardiac traits in South Asian and Chinese populations. Furthermore, the addition of a small and individually underpowered Chinese ancestry-specific GWAS (Chinese) can improve the predictive performance of PGS. This lends hope that performing modestly sized ancestry-specific GWAS could be sufficient to generate PGS with comparable performance to European ancestry populations^16^.

In conclusion, this study identifies multiple clinical applications for a PGS in HCM, including in general population screening, stratifying of rare variant carriers into higher and lower risk of penetrant HCM, and as a novel risk predictor of adverse outcomes in individuals with HCM.

## Online Methods

### GWAS meta-analysis and multi-trait analysis GWAS

The base data for the HCM PGS is from the largest HCM GWAS, consisting of 5900 cases and 68359 unrelated controls, from 7 cohorts (100,000 Genomes Project [GeL], BioResource Rare Diseases [BRRD], HCM Registry [HCMR], and clinical cohorts from Canada, Italy, Netherlands, and the UK)^10^. HCM was defined as primary left ventricular hypertrophy, in the absence of secondary causes (uncontrolled hypertension, aortic valve disease, infiltrative cardiomyopathic processes, and cases arising from complex syndromes), using a combination of clinical, imaging and ICD-code definitions. Detailed information on cohorts included in the GWAS are provided in the original publication^10^. Cases and controls included in the HCM GWAS were of European ancestry. Leveraging the increased power generated from jointly analyzing genetically-correlated traits using the multi-trait analysis of GWAS (MTAG) method^30^, MTAG of HCM was performed using *mtag*^30^ with three genetically correlated traits (LV concentricity, LV end systolic volume and LV circumferential strain)^10^.

### Polygenic score derivation and evaluation

Individual SNP weighted scores were generated from the primary discovery GWAS and MTAG. The base GWAS and MTAG summary statistics were filtered to exclude rare and uncommon variants (minor allele frequency [MAF] <1%), and ambiguous SNPs that were not resolvable by strand flipping. A locus on chromosome 11 surrounding *MYBPC3* was found to be associated with HCM in only sarcomere-positive HCM, specifically in one cohort (NL) and was determined to represent a founder effect^10^. Variants with P<1×10^−5^ on chromosome 11 from 30000000 to 80000000 (GRCh37) were excluded from PGS calculation.

We calculated HCM PGS for unrelated (3^rd^ degree or closer), White British participants in the UK Biobank (UKB; application number 47602) using variants that passed genotyping QC (MAF>1%, genotyping rate >0.99, HWE 1×10^−6^). Variants overlapping the base, target, and LD reference set (1000 Genomes Project Phase 3 European ancestry) were included. The individual SNP scores were generated using *PRS-CS*, a package that uses a Bayesian framework to model linkage disequilibrium (LD) using an external LD reference set and a continuous shrinkage prior on SNP effect sizes^31^. The phi constant was automatically selected by *PRS-CS* in an unsupervised approach (PRS-CS auto). Whole genome PGS scores for all included UKB individuals and testing cohorts were calculated using the *–score* function in *PLINKv1*.*9*^32^.

PGS was applied and tested within a range of cohorts and clinical settings. Given that a key factor in the predictive power of PGS is the power of the base GWAS^33^, we first compared the performance of PGS generated using GWAS (PGS_GWAS_, 376,730 SNP predictors) and MTAG^10^ (PGS_MTAG_, 374,113 SNP predictors) summary statistics in 343,182 unrelated White British ancestry participants in the UKB. Predictive performance of PGS was assessed by comparing Nagelkere’s R2, area under the receiver operating characteristic (AUROC), and association with HCM (OR per PGS standard deviation).

Inclusion of participants in both the testing and GWAS datasets results in substantial inflation of PGS performance^34^. To prevent this, where case-control PGS testing was performed in a cohort that was included in the main GWAS (for example, GeL), PGS was generated using a leave-one-study-out GWAS and MTAG that did not include the cohort. All other methods for PGS generation remained the same. In situations where only cases are included in the assessment of PGS, the overall MTAG results were used.

### Cohorts

#### UK Biobank

UKB is a population-based cohort study from half a million UK participants, with detailed clinical, imaging, and genetic data. Participants from UKB that were included in testing were unrelated (3^rd^ degree or closer) and of White British ancestry. HCM cases were identified from self-report clinical data (hospital admissions and death registry), and CMR imaging (LV maximum wall thickness >15mm). Time to clinical event was identified from UKB first occurrences data, operation dates, and death dates. Participants in the imaging substudy were randomly invited from the overall cohort. Each underwent CMR at 1.5-T. Segmentation of the cine images was performed by using a deep learning neural network algorithm and have previously been reported^6^.

#### 100,000 Genomes Project

The 100,000 Genomes Project (GeL) is a national UK programme that recruited probands with rare diseases and cancer from clinical centres, together with family members, and performed germline and somatic (for a subset of participants with cancer) whole genome sequencing^17,35^. HCM cases were identified from HPO terms at time of study recruitment, and ICD codes from preceding and subsequent clinical episodes.

#### Erasmus Medical Centre cohort

To evaluate the role of PGS in modulating penetrance of sarcomeric variants in relatives of HCM cases we used a subset of 214 relatives of HCM probands from an ongoing HCM registry at the Erasmus Medical Centre (EMC, Rotterdam)^36,37^. All individuals were carriers of pathogenic sarcomeric variants, with the exclusion of homozygous carriers or those carrying multiple pathogenic or likely pathogenic variants.

#### Royal Brompton and Harefield Hospitals cohort

Unrelated White British HCM cases from the Royal Brompton & Harefield Hospitals Cardiovascular Research Biobank^38^ (RBH) were used to assess the effect of PGS on CMR imaging traits. Data from the clinical CMR scan taken at or prior to study recruitment were used, and where sequential CMR scans were available, follow-up imaging data was recorded to identify changes in imaging traits.

### Statistical analysis

In the UKB, PGS model performance was assessed using Nagelkerke’s R^2^, adjusting the null model for age, age^2^, sex and first ten principal components. The predictive AUROC was determined using a randomly subsetted training (70%) and validation (30%) cohort using R-package *pROC* (v1.18.0)^39^. For association between PGS and HCM status in UKB and GeL, logistic regression was performed adjusting for age, age^2^, sex and first ten principal components. In EMC, this was assessed using Wald logistic mixed-effects model using *GMMAT* (v1.3.2) adjusting for fixed-effects of sex, age, age^2^ and first four principal components and incorporating a genetic relatedness matrix (GRM) estimated using GCTA (v1.92.2beta)^40^ as a random effect. For quantitative imaging traits in UKB and RBH, PGS association was evaluated using linear regression adjusting for age, age^2^, sex, first ten principal components, SBP and BSA and differences between means in stratified groups was performed with ANCOVA testing adjusted for age, age^2^, sex, BSA, SBP and first ten principal components. In EMC, the association of PGS with LV maximum wall thickness was assessed using a linear mixed-effects model using *coxme* (v2.2-17)^41^, adjusting for sex, age at imaging, age at imaging^2^, imaging modality (CMR vs. TTE), first four principal components and the GRM. Time-to-event data in UKB was evaluated using cox-proportional hazards model, adjusting for age, age^2^, sex and first ten principal components using *survival* (v2.44-1.1). Hazards assumption for proportionality was assessed, and for outcomes that did not include death, competing risk analysis was performed. In EMC, the association between PGS and clinical events was assessed using a Cox proportional hazards mixed-effects model using R-package *coxme* (v2.2-17)^41^ adjusted for sex, first four principal components, GRM and presence of *MYH7* rare variant genotype status. All survival curves were created using *survminer* (v0.4.9). Although clinical data was complete for most individuals in all cohorts, where missing data was present, individuals were excluded from analysis. All statistical analysis was performed in R. For multi-ancestry analysis, ancestry as categorical variable was included in the regression model. Was included in the regression model.

### Rare variant status

The pathogenicity of rare variants in 8 definitive HCM-causing genes^42^ (*MYBPC3, MYH7, TNNT2, TNNI3, TPM1, ACTC1, MYL3*, and *MYL2*) was determined using broadly similar approaches across cohorts in line with ACMG guidelines^43^ (Supplementary Methods). Individuals without pathogenic or likely pathogenic variants were identified as gene negative.

### Outcomes

For a diagnosis of HCM in the UKB, HCM cases were identified from self-reporting, ICD codes from hospital encounters and the national death register, and CMR imaging (maximum LV wall thickness >15mm), in the absence of aortic stenosis (Supplementary Methods). For the analysis of imaging traits in HCM cases, we further refined the diagnosis by restricting only to individuals with a LV maximum wall thickness of at least 13mm. PGS association with a range of HCM-relevant cardiac imaging traits associated with cardiac structure (maxLVWT, LVEDV, LVESV, LA volume and fractal dimensions) and function (LVEF, and strain measurements) was tested. Longitudinal risk of time to HCM diagnosis and for major adverse cardiovascular events was assessed. Clinical and operative outcomes were selected based on their relevance to HCM, incorporating self-reported diagnoses, hospital admission events, primary care records, death records (Supplementary Methods).

Diagnosis of HCM in additional cohorts (EMC, GeL and RBH) and clinical outcomes in EMC are reported in Supplementary Methods.

### PGS generation and testing in diverse ancestry groups

PGS generated using European ancestry GWAS have weaker performance when tested in more diverse ancestry populations^12-15^. We first aimed to evaluate PGS performance in participants of Afro-Caribbean (n=661), East Asian (n=504) and South Asian (n=489) ancestry groups in UKB, by applying ancestry-specific 1000 Genomes Project LD reference sets to the European ancestry GWAS and MTAG when generating PGS. Ancestries of UKB participants was determined based on self-reported ancestry, followed by visualization of principal component plots and manual selection of principal component thresholds. Given that PGS are not comparable between differing ancestries due to underlying differing genetic architecture, analyses using PGS as a continuous variable were restricted within single ancestry groups. For analysis stratifying by quantiles, quantiling was first performed within each ancestry before being combined with other ancestries.

PRS-CSx extends the Bayesian polygenic modeling and prediction methods of PRS-CS by combining GWAS summary statistics from multiple ancestry groups and has been shown to improve cross ancestry prediction^16^. We aimed to evaluate the performance of PGS generated using this approach for the prediction of HCM-associated CMR traits in East Asian ancestry participants in the UKB, by combining the European ancestry GWAS with a small Chinese ancestry GWAS (Singapore cohort).

### Singapore HCM GWAS

GWAS was performed of 184 cases and 776 controls of Chinese ancestry. Genotyping was performed using Infinium OmniExpress-24 kit (Illumina). Imputation was performed on the Michigan Imputation Server^44^ using Minimac4 (version 1.5.7) and East Asian reference genomes (1000 Genomes Phase 3 version 5^45^, 1000 Genomes Phase 1 version 3^45^ and Genome Asia Pilot^46^). Post imputation QC was performed at variant (HWE >1×10^−7^, genotyping >0.95, information score >0.5 and MAF >1%) level. Chinese ancestry individuals were identified using principal component analysis and one of a pair of 2^nd^ degree or closer relatives was retained. GWAS was tested using SNPTEST^47^ adjusting for age, sex and first ten principal components.

### Phenome-wide association study and Mendelian randomization

PheWAS was performed in the UKB to investigate pleiotropic effects of the HCM PGS. ICD-9 and ICD-10 codes from death records and hospital admission episodes were translated to Phecodes (Phecode Map 1.2)^48,49^. For phenotypes with at least 20 cases, PGS-phenotype association was tested using logistic regression adjusted for age, age^2^, sex and first ten principal components as covariates. Significance threshold was adjusted for the total number of phenotypes tested (P<2.72×10^−5^), and data presented with Manhattan plots grouping by body system. PheWAS was performed using the *PheWAS* package^50^ in R version 4.0.3.

To further evaluate directionality of effect for key significant PheWAS associations (hypertension, dyslipidemia, type 2 diabetes), two-sample bidirectional Mendelian randomization (MR) was performed for relevant quantitative traits (systolic and diastolic blood pressure, hypercholesterolemia, glycated hemoglobin, and body mass index). To maximize MR power, the exposure trait GWAS with the largest number of significant SNPs after harmonization from the IEU GWAS database was used as the instrument (Table S3)^51-54^, and was harmonized with the HCM MTAG. MR Steiger directionality test was used to evaluate the causal direction between exposure and HCM. Two-sample MR using MR Egger regression was performed using the *TwoSampleMR*^*55,56*^ and *MRInstruments* packages in R version 4.0.3.

## Data Availability

All data produced in the present study are available upon reasonable request to the authors. PGS scores will be made available on the Polygenic Score Catalog (www.pgscatalog.com) upon publication following peer-review of the article.

## Ethics declaration

All patients gave written informed consent, and all studies were approved by the relevant regional research ethics committees, and adhered to the principles set out in the Declaration of Helsinki. The UK Biobank study was reviewed by the National Research Ethics Service (11/NW/0382, 21/NW/0157). The 100,000 Genomes Project was reviewed by the National Research Ethics Service (14/EE/1112 and 13/EE/032). The Royal Brompton Biobank was reviewed and approved by the South Central – Hampshire B Research Ethics Committee (09/H0504/104+5 and 19/SC/0257). The Erasmus Medical Center was reviewed and approved by the Erasmus MC Medical Ethical Review Committee. All Singaporean participants recruited from the National Heart Center Singapore gave written informed consent and the study was approved by the Singhealth Centralised Institutional Review Board (2020/2353) and the Singhealth Biobank Research Scientific Advisory Executive Committee (SBRSA 2019/001v1).

## Data and code availability

Data from UK Biobank can be requested from the UK Biobank Access Management System. Data from 100,000 Genomes Project can be accessed following application to join the Genomics England Clinical Interpretation Partnership. The PGS generated using GWAS and MTAG summary statistics will be made available on the Polygenic Score Catalog (www.pgscatalog.org) upon publication following peer-review of the article. GWAS and

MTAG summary statistics will be made available in the GWAS Catalog (www.ebi.ac.uk/gwas) upon publication following peer-review of the associated article^10^.The analyses reported in this article rely on previously published software, as detailed in the Methods, and custom code will be made available upon request.

## Acknowledgements and funding sources

This work was supported by funding from the British Heart Foundation [RE/18/4/34215, FS/IPBSRF/22/27059, FS/15/81/31817, FS/ICRF/21/26019, RG/19/6/34387, RE/13/1/30181, BBC/F/21/220106]; the Medical Research Council [MC_UP_1605/13]; Wellcome Trust [107469/Z/15/Z]; the National Institute for Health Research (NIHR) Imperial College Biomedical Research Centre; NIHR Royal Brompton Cardiovascular Biomedical Research Unit; Sir Jules Thorn Charitable Trust [21JTA]; National Heart Lung Institute Foundation; Royston Centre for Cardiomyopathy Research; the Dutch Heart Foundation [03-007-2022-0035, CVON PRIME]; Rosetrees Trust; EJP-RD, Leducq Foundation [17CVD02]; Amsterdam Cardiovascular Sciences; European Commission [LSHM-CT-2007-037273, HEALTH-F2-2013-601456]; European Joint Programme Rare Diseases [LQTS-NEXT, ZonMW project 40-46300-98-19009].

This research has been conducted in part using the UK Biobank Resource under Application Numbers 18545, 40616 and 47602. This research was made possible through access to the data and findings generated by the 100,000 Genomes Project under Project Number 667.

The 100,000 Genomes Project is managed by Genomics England Limited (a wholly owned company of the Department of Health and Social Care), and funded by the National Institute for Health Research and NHS England. The Wellcome Trust, Cancer Research UK and the Medical Research Council have also funded research infrastructure. The 100,000 Genomes Project uses data provided by patients and collected by the National Health Service as part of their care and support. Genotyping was supported by the Institute of Psychiatry Psychology and Neuroscience (IoPPN) Genomics & Biomarker Core Facility within King’s College London, who gratefully acknowledge capital equipment funding from the Maudsley Charity [980] and Guy’s and St Thomas’s Charity [STR130505].

The views expressed in this work are those of the authors and not necessarily those of the funders. For the purpose of open access, the authors have applied a Creative Commons Attribution (CC BY) licence to any Author Accepted Manuscript version arising from this submission.

## Notes

### Competing Interest Statement

The authors have declared no competing interest.

### Author Declarations

All patients gave written informed consent, and all studies were approved by the relevant regional research ethics committees, and adhered to the principles set out in the Declaration of Helsinki. The UK Biobank study was reviewed by the National Research Ethics Service (11/NW/0382, 21/NW/0157). The 100,000 Genomes Project was reviewed by the National Research Ethics Service (14/EE/1112 and 13/EE/032). The Royal Brompton Biobank was reviewed and approved by the South Central - Hampshire B Research Ethics Committee (09/H0504/104+5 and 19/SC/0257). The Erasmus Medical Center was reviewed and approved by the Erasmus MC Medical Ethical Review Committee. All Singaporean participants recruited from the National Heart Center Singapore gave written informed consent and the study was approved by the Singhealth Centralised Institutional Review Board (2020/2353) and the Singhealth Biobank Research Scientific Advisory Executive Committee (SBRSA 2019/001v1).

## References

1. Semsarian, C., Ingles, J., Maron, M.S. & Maron, B.J. New perspectives on the prevalence of hypertrophic cardiomyopathy. J Am Coll Cardiol 65, 1249–1254 (2015).

2. Ommen, S.R., et al. 2020 AHA/ACC Guideline for the Diagnosis and Treatment of Patients With Hypertrophic Cardiomyopathy: A Report of the American College of Cardiology/American Heart Association Joint Committee on Clinical Practice Guidelines. Circulation 142, e558–e631 (2020).

3. Elliott, P.M., et al. 2014 ESC Guidelines on diagnosis and management of hypertrophic cardiomyopathy: the Task Force for the Diagnosis and Management of Hypertrophic Cardiomyopathy of the European Society of Cardiology (ESC). Eur Heart J 35, 2733–2779 (2014).

4. Marian, A.J. & Braunwald, E. Hypertrophic Cardiomyopathy: Genetics, Pathogenesis, Clinical Manifestations, Diagnosis, and Therapy. Circ Res 121, 749–770 (2017).

5. McKenna, W.J. & Judge, D.P. Epidemiology of the inherited cardiomyopathies. Nature Reviews Cardiology 18, 22–36 (2021).

6. de Marvao, A., et al. Phenotypic Expression and Outcomes in Individuals With Rare Genetic Variants of Hypertrophic Cardiomyopathy. J Am Coll Cardiol 78, 1097–1110 (2021).

7. Jurgens, S.J., et al. Analysis of rare genetic variation underlying cardiometabolic diseases and traits among 200,000 individuals in the UK Biobank. Nature Genetics 54, 240–250 (2022).

8. Harper, A.R., et al. Common genetic variants and modifiable risk factors underpin hypertrophic cardiomyopathy susceptibility and expressivity. Nat Genet 53, 135–142 (2021).

9. Tadros, R., et al. Shared genetic pathways contribute to risk of hypertrophic and dilated cardiomyopathies with opposite directions of effect. Nat Genet 53, 128–134 (2021).

10. Tadros, R., et al. Large scale genome-wide association analyses identify novel genetic loci and mechanisms in hypertrophic cardiomyopathy. medRxiv, 2023.2001.2028.23285147 (2023).

11. Biddinger, K.J., et al. Rare and Common Genetic Variation Underlying the Risk of Hypertrophic Cardiomyopathy in a National Biobank. JAMA Cardiol 7, 715–722 (2022).

12. Thompson, D.J., et al. UK Biobank release and systematic evaluation of optimised polygenic risk scores for 53 diseases and quantitative traits. medRxiv, 2022.2006.2016.22276246 (2022).

13. Duncan, L., et al. Analysis of polygenic risk score usage and performance in diverse human populations. Nat Commun 10, 3328 (2019).

14. Martin, A.R., et al. Clinical use of current polygenic risk scores may exacerbate health disparities. Nat Genet 51, 584–591 (2019).

15. Kamiza, A.B., et al. Transferability of genetic risk scores in African populations. Nature Medicine 28, 1163–1166 (2022).

16. Ruan, Y., et al. Improving polygenic prediction in ancestrally diverse populations. Nature Genetics 54, 573–580 (2022).

17. Caulfield, M., et al. The National Genomics Research and Healthcare Knowledgebase. (figshare, 2019).

18. Khera, A.V., et al. Genome-wide polygenic scores for common diseases identify individuals with risk equivalent to monogenic mutations. Nat Genet 50, 1219–1224 (2018).

19. Mars, N., et al. The role of polygenic risk and susceptibility genes in breast cancer over the course of life. Nat Commun 11, 6383 (2020).

20. Ingles, J., et al. Nonfamilial Hypertrophic Cardiomyopathy: Prevalence, Natural History, and Clinical Implications. Circ Cardiovasc Genet 10 (2017).

21. Watkins, H. Time to Think Differently About Sarcomere-Negative Hypertrophic Cardiomyopathy. Circulation 143, 2415–2417 (2021).

22. Ormondroyd, E., et al. Secondary findings in inherited heart conditions: a genotype-first feasibility study to assess phenotype, behavioural and psychosocial outcomes. European Journal of Human Genetics 28, 1486–1496 (2020).

23. Adeyemo, A., et al. Responsible use of polygenic risk scores in the clinic: potential benefits, risks and gaps. Nature Medicine 27, 1876–1884 (2021).

24. Spirito, P., et al. Clinical course and prognosis of hypertrophic cardiomyopathy in an outpatient population. N Engl J Med 320, 749–755 (1989).

25. Weissler-Snir, A., Dorian, P., Rakowski, H., Care, M. & Spears, D. Primary prevention implantable cardioverter-defibrillators in hypertrophic cardiomyopathy-Are there predictors of appropriate therapy? Heart Rhythm 18, 63–70 (2021).

26. Maron, B.J., et al. Prevalence of hypertrophic cardiomyopathy in a general population of young adults. Echocardiographic analysis of 4111 subjects in the CARDIA Study. Coronary Artery Risk Development in (Young) Adults. Circulation 92, 785–789 (1995).

27. Maron, B.J., et al. Prevalence of hypertrophic cardiomyopathy in a population-based sample of American Indians aged 51 to 77 years (the Strong Heart Study). Am J Cardiol 93, 1510–1514 (2004).

28. Eberly, L.A., et al. Association of Race With Disease Expression and Clinical Outcomes Among Patients With Hypertrophic Cardiomyopathy. JAMA Cardiology 5, 83–91 (2020).

29. Bai, Y., et al. Prevalence, incidence and mortality of hypertrophic cardiomyopathy based on a population cohort of 21.9 million in China. Scientific Reports 12, 18799 (2022).

30. Turley, P., et al. Multi-trait analysis of genome-wide association summary statistics using MTAG. Nature Genetics 50, 229–237 (2018).

31. Ge, T., Chen, C.Y., Ni, Y., Feng, Y.A. & Smoller, J.W. Polygenic prediction via Bayesian regression and continuous shrinkage priors. Nat Commun 10, 1776 (2019).

32. Purcell, S., et al. PLINK: a tool set for whole-genome association and population-based linkage analyses. Am J Hum Genet 81, 559–575 (2007).

33. Dudbridge, F. Polygenic Epidemiology. Genet Epidemiol 40, 268–272 (2016).

34. Wray, N.R., et al. Pitfalls of predicting complex traits from SNPs. Nat Rev Genet 14, 507–515 (2013).

35. Smedley, D., et al. 100,000 Genomes Pilot on Rare-Disease Diagnosis in Health Care - Preliminary Report. N Engl J Med 385, 1868–1880 (2021).

36. Vriesendorp, P.A., et al. Long-term outcomes after medical and invasive treatment in patients with hypertrophic cardiomyopathy. JACC. Heart failure 2 6, 630–636 (2014).

37. van Velzen, H.G., et al. Effect of Gender and Genetic Mutations on Outcomes in Patients With Hypertrophic Cardiomyopathy. Am J Cardiol 122, 1947–1954 (2018).

38. Walsh, R., et al. Reassessment of Mendelian gene pathogenicity using 7,855 cardiomyopathy cases and 60,706 reference samples. Genet Med 19, 192–203 (2017).

39. Robin, X., et al. pROC: an open-source package for R and S+ to analyze and compare ROC curves. BMC Bioinformatics 12, 77 (2011).

40. Yang, J., Lee, S.H., Goddard, M.E. & Visscher, P.M. GCTA: a tool for genome-wide complex trait analysis. Am J Hum Genet 88, 76–82 (2011).

41. Ripatti, S. & Palmgren, J. Estimation of multivariate frailty models using penalized partial likelihood. Biometrics 56, 1016–1022 (2000).

42. Ingles, J., et al. Evaluating the Clinical Validity of Hypertrophic Cardiomyopathy Genes. Circ Genom Precis Med 12, e002460 (2019).

43. Richards, S., et al. Standards and guidelines for the interpretation of sequence variants: a joint consensus recommendation of the American College of Medical Genetics and Genomics and the Association for Molecular Pathology. Genet Med 17, 405–424 (2015).

44. Das, S., et al. Next-generation genotype imputation service and methods. Nat Genet 48, 1284–1287 (2016).

45. Auton, A., et al. A global reference for human genetic variation. Nature 526, 68–74 (2015).

46. Wall, J.D., et al. The GenomeAsia 100K Project enables genetic discoveries across Asia. Nature 576, 106–111 (2019).

47. Marchini, J., Howie, B., Myers, S., McVean, G. & Donnelly, P. A new multipoint method for genome-wide association studies by imputation of genotypes. Nature Genetics 39, 906–913 (2007).

48. Wei, W.-Q., et al. Evaluating phecodes, clinical classification software, and ICD-9-CM codes for phenome-wide association studies in the electronic health record. in PLoS One, Vol. 12 e0175508 (2017).

49. Wu, P., et al. Mapping ICD-10 and ICD-10-CM Codes to Phecodes: Workflow Development and Initial Evaluation. JMIR Med Inform 7, e14325 (2019).

50. Carroll, R.J., Bastarache, L. & Denny, J.C. R PheWAS: data analysis and plotting tools for phenome-wide association studies in the R environment. Bioinformatics 30, 2375–2376 (2014).

51. Evangelou, E., et al. Genetic analysis of over 1 million people identifies 535 new loci associated with blood pressure traits. Nat Genet 50, 1412–1425 (2018).

52. Elsworth, B., et al. The MRC IEU OpenGWAS data infrastructure. bioRxiv, 2020.2008.2010.244293 (2020).

53. Chen, J., et al. The trans-ancestral genomic architecture of glycemic traits. Nat Genet 53, 840–860 (2021).

54. Willer, C.J., et al. Discovery and refinement of loci associated with lipid levels. Nat Genet 45, 1274–1283 (2013).

55. Hemani, G., Tilling, K. & Davey Smith, G. Orienting the causal relationship between imprecisely measured traits using GWAS summary data. PLoS Genet 13, e1007081 (2017).

56. Hemani, G., et al. The MR-Base platform supports systematic causal inference across the human phenome. Elife 7(2018).

