## Supplementary Figures for "Evaluation of polygenic score for hypertrophic cardiomyopathy in the general population and across clinical settings"

**Figure S1****: PGS and cohort overview**

**Figure S2****: PGS in the general population**

**Figure S3****: HCM PGS and cardiac imaging traits in the general population**

**Figure S4****: PGS in non-White ancestry groups**

**Figure S5****: Singapore Chinese ancestry HCM case-control GWAS**

**Figure S6****: PGS in sarcomere-positive carriers**

**Figure S7****: PGS in SARC-PLP carrier relatives of HCM cases**

**Figure S8****: PGS predicts adverse events and disease severity in HCM cases**

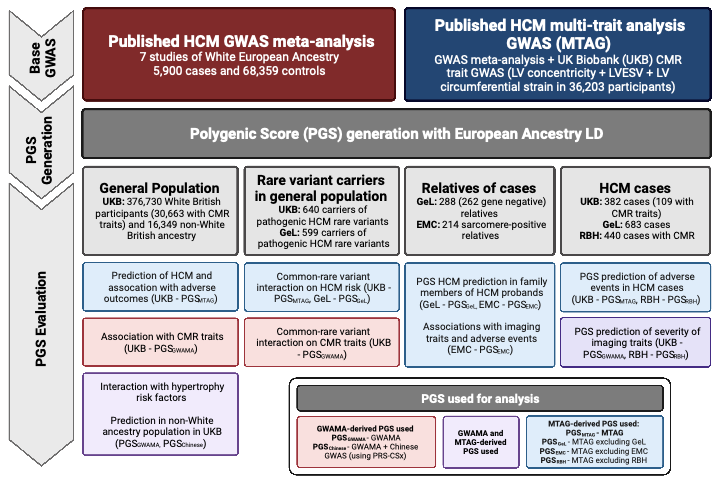

**Figure S1: PGS and cohort overview.** Bayesian genome-wide PGS were generated from a published European-ancestry hypertrophic cardiomyopathy (HCM) GWAS meta-analysis of seven case-control studies (comprising 5,900 cases and 68,359 controls; PGS_GWAS_), and multi-trait analysis of GWAS (analysing HCM with three genetically-correlated quantitative traits measured using cardiac MRI [CMR] in 36,203 UKB participants: LV concentricity, LV end systolic volume and LV circumferential strain; PGS_MTAG_). In order to minimise inflation due to overlap of samples in the base GWAS and cohort in which the PGS is being evaluated, leave-one-study-out GWAS meta-analyses were performed to generate base GWAS without any sample overlap. For example, when PGS was evaluated in GeL, first a GWAS meta-analysis excluding the GeL cohort was performed, which was then used to generate PGS that was tested in GeL. Similarly, for association of CMR traits in the UKB, given that the MTAG used GWAS summary statistics of imaging traits performed in the UKB, PGS derived from GWAS meta-analysis (PGS_GWAMA_) was used rather than PGS derived from MTAG. All PGS performed similarly well in their associations with population risks of HCM in the UKB (Table S1). UKB – UK Biobank, GeL – 100,000 Genomes Project; EMC – Erasmus Medical Centre, Netherlands; RBH – Royal Brompton Hospital, UK. LV – left ventricle/ventricular, LVESV – LV end-systolic volume.

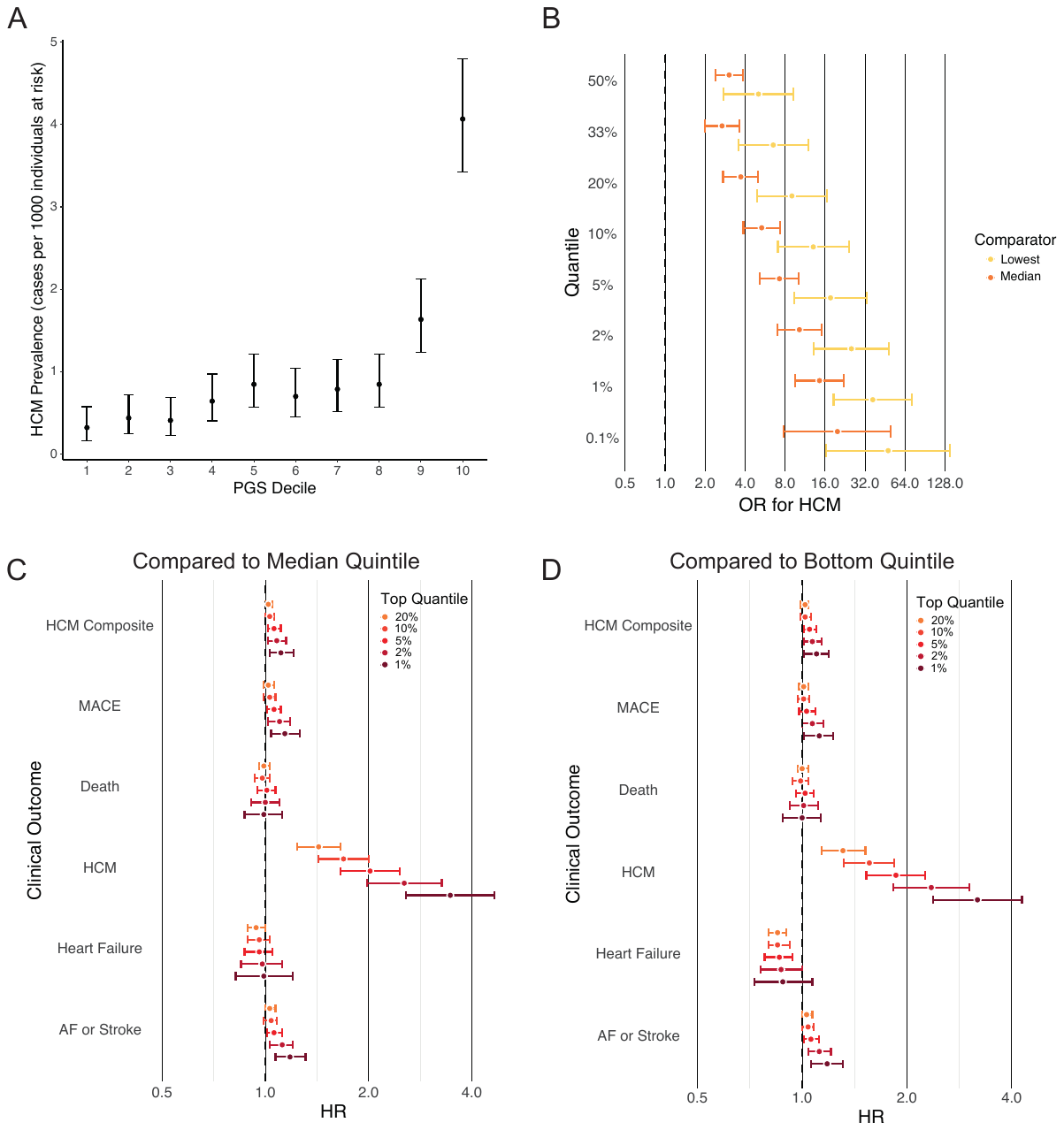

**Figure S2: PGS in the general population.** (A) Prevalence of HCM in UKB in each decile. (B) OR for HCM comparing a range of top quantiles with median 20% and lowest quantiles. (C and D) Hazards ratio for adverse cardiovascular events when comparing with the median (C) and bottom (D) quintiles. HCM composite consists of death, heart failure, AF, stroke, cardiac arrest, septal reduction therapy (surgical myectomy or alcohol septal ablation), ICD implantation, LVAD implantation, or cardiac transplantation. MACE consists of HCM diagnosis, heart failure, AF, stroke or cardiac arrest.

A B
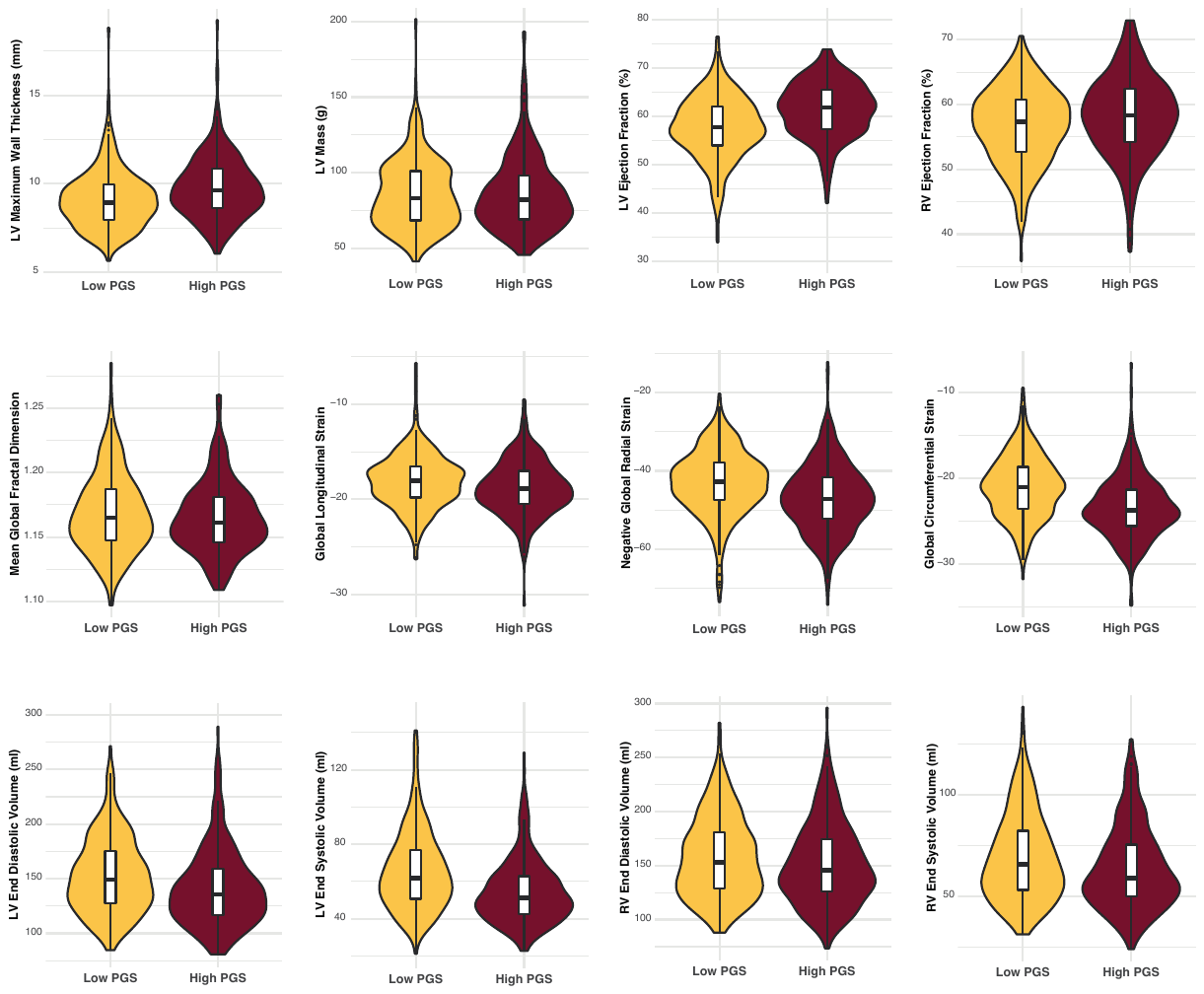

**Figure S3: HCM PGS and cardiac imaging traits in the general population.** (A) PGS_GWAMA_ associations with machine-learning derived quantitative CMR traits in 30,663 unrelated participants in the UKB. Univariate regression line of PGS and trait (blue line). (B) Violin plot of quantitative CMR traits in top and bottom PGS centiles. Box plots indicate mean and standard deviation. LV: left ventricle.

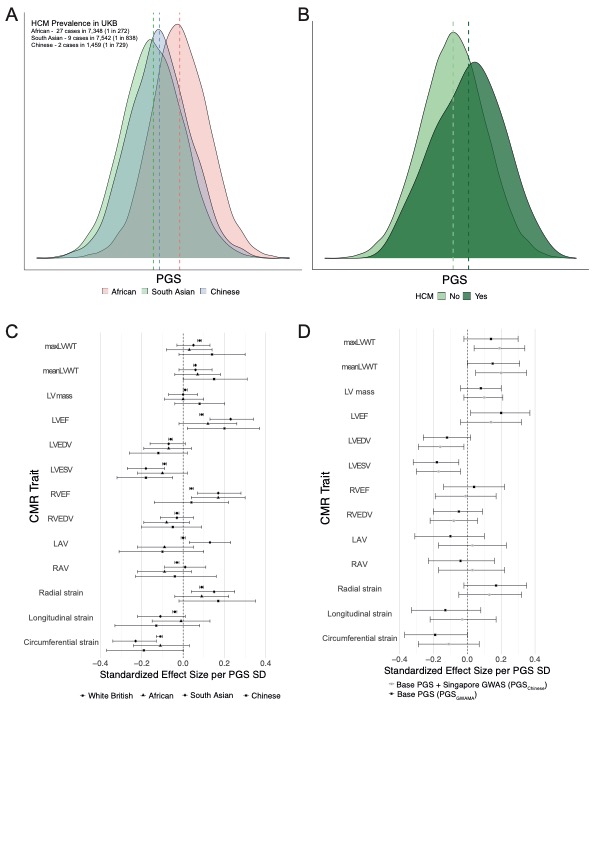
**Figure S4: PGS in non-White ancestry groups.** (A) PGS distribution across ancestries in South Asian (7,538 participants), Afro-Caribbean (7,346) and East Asian (1,457) participants in the UKB, and in non-White British case and controls. (B) Pooled PGS distribution in non-White ancestry individuals in UKB, stratified by case-control status. (C) Standardised effect size per PGS SD on CMR traits in non-White ancestry groups. (D) Standardised effect size per PGS SD on CMR traits in 1,457 Chinese ancestry individuals in UKB using standard PGS (PGS_GWAMA_) and PGS generated with the addition of Singapore GWAS using PGS-CSx (PGS_Chinese_).

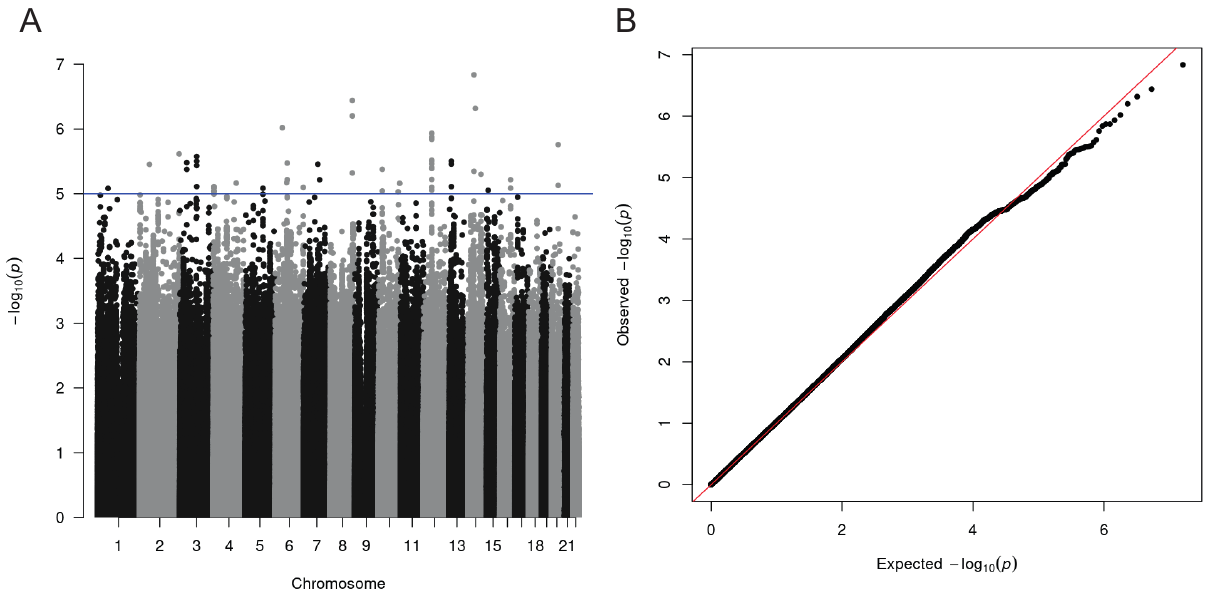

**Figure S5: Singapore Chinese ancestry HCM case-control GWAS.** (A) Manhattan and (B) QQ plot of Singapore HCM case-control GWAS consisting of 174 HCM cases and 776 controls, all of Chinese ancestry (lambda GC 1.03). Blue line indicates suggestive significance threshold (P<1x10^-5^). No SNPs reached genome-wide significance (P<5x10^-8^).

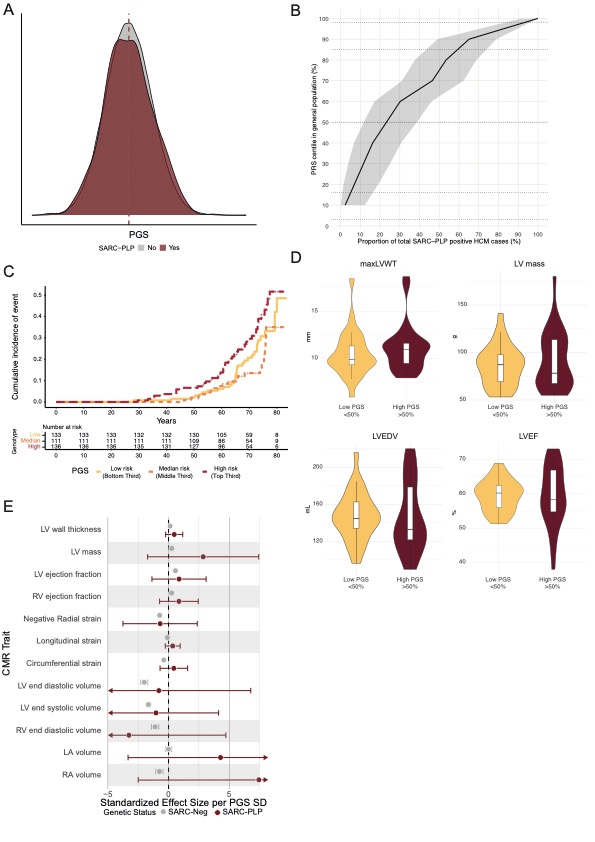

**Figure S6: PGS in sarcomere-positive carriers.** (A) PGS distribution in 318,945 whole-exome sequenced UKB participants with (n=640) and without (n=318,305) pathogenic HCM-causing variants shows no difference between groups, suggesting again any marked selection or suvival bias. (B) Cumulative curve of gene positive HCM cases across PGS centiles. Dashed lines represent population mean, ±1 SD and ±2 SD. (C) Time to HCM composite outcome (comprising death, cardiac arrest, atrial fibrillation, stroke, HCM, heart failure, ICD implantation, septal reduction therapy, LVAD implantation and cardiac transplantation) in top, median and bottom quintiles. (E) Standardized effect size of PGS SD on quantitative CMR traits known to be affected in HCM, in those with (N=53) and without (N=29,002) SARC-PLP variants highlighting directionally concordant trends in both groups.

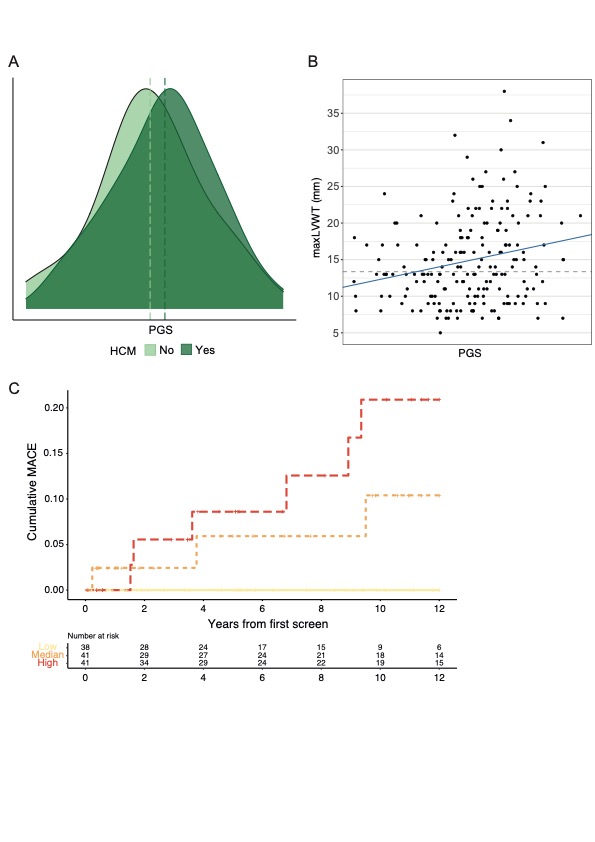

**Figure S7: PGS in SARC-PLP carrier relatives of HCM cases.** (A) PGS distribution in 214 relatives of HCM cases in the Erasmus cohort, stratified by HCM status. (B) Scatter plot of maxLVWT against PGS_EMC_ among 194 sarcomere-positive relatives of HCM index patients from Erasmus cohort. Univariate linear regression line (blue line) and common diagnostic cutoff for HCM in relatives of cases (13 mm, grey dotted line). (C) Cumulative major adverse cardiovascular events (MACE) after initial screening in sarcomere-positive relatives of HCM probands stratified by PGS_EMC_ in the top, middle and bottom quintiles. MACE was defined as a composite of septal reduction therapy, cardiac transplantation, aborted cardiac arrest, appropriate defibrillator shock, or sudden cardiac death.

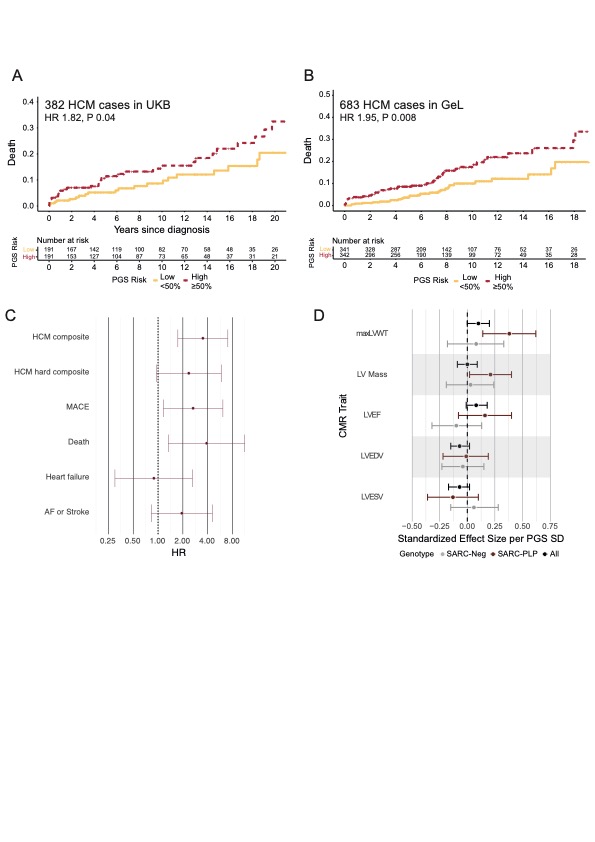
**Figure S8: PGS predicts adverse events and disease severity in HCM cases**. Cumulative hazard curves for all-cause mortality in UKB (n=382) (A) and GeL (n=683) (B) after HCM diagnosis, stratified by median PGS. (C) Risk of adverse events in HCM cases, comparing top and bottom PGS quintiles in UKB HCM cases. (D) Effect of PGS on CMR imaging traits in 440 HCM cases at the Royal Brompton Hospital, stratified by genetic status (101 SARC-PLP, 104 SARC-Neg) and all (440 cases, including 235 with VUS or unknown variant status). HCM composite adverse outcome consists of death, heart failure, atrial fibrillation, stroke, cardiac arrest, septal reduction therapy (myectomy or alcohol septal ablation), ICD implantation, LVAD implantation or cardiac transplantation. HCM hard composite outcome consists of death, stroke, cardiac arrest, myectomy, LVAD implantation or cardiac transplantation. Major adverse cardiovascular events (MACE) consists of heart failure, atrial fibrillation, stroke or cardiac arrest.
