## Supplementary Methods for "Evaluation of polygenic score for hypertrophic cardiomyopathy in the general population and across clinical settings"

HCM definitions across cohorts

Clinical outcomes in UK Biobank

EMC cohort methods

Variant pathogenicity

Supplementary References

**HCM definition across cohorts**

*UK Biobank*

- Self-reported hypertrophic cardiomyopathy (HCM) or hypertrophic obstructive cardiomyopathy (HOCM) at time of study enrollment (UKB field 20002).
- ICD-10 (I42.1, I42.2) or ICD-9 (425.1) code for HCM or HOCM recorded on hospital episode statistic or on death record (<https://github.com/UK-Digital-Heart-Project/UKBB_filter>), in the absence of aortic stenosis (UKB fields 40001, 40002, 41202, 41204, and 41270).
- Maximum LV wall thickness of at least 15mm in the absence of aortic stenosis with SBP<140mmHg on date of scan, quantified using approach in *De Marvao et al.^1^*

*100K Genomes Project*

- Referral to study with diagnosis of HCM or HOCM from HPO terms.
- ICD-10 (I42.1, I42.2) code for HCM or HOCM recorded on hospital episode statistic.

*Erasmus Medical Centre, NL*

- Clinical diagnosis of HCM/HOCM using guideline-based criteria (imaging, personal and family history, examination, genetic, and other investigations)^2,3^.

*Royal Brompton Hospital, UK*

- Clinical diagnosis of HCM/HOCM using guideline-based criteria (imaging, personal and family history, examination, genetic, and other investigations)^2,3^.

*National Heart Center Singapore, Singapore*

- Clinical diagnosis of HCM/HOCM using guideline-based criteria (imaging, personal and family history, examination, genetic, and other investigations)^2,3^.

**Clinical Outcomes in UK Biobank**

HCM composite:

- Death
- HCM
- Heart failure
- Atrial fibrillation/flutter
- Stroke
- Cardiac arrest
- Septal reduction therapy (surgical myectomy or alcohol septal ablation)
- ICD implantation
- LVAD implantation
- Cardiac transplantation

HCM hard composite:

- Death
- Stroke
- Cardiac arrest
- Surgical myectomy
- LVAD implantation
- Cardiac transplantation

Major adverse cardiovascular:

- HCM
- Heart failure
- Atrial fibrillation/flutter
- Stroke
- Cardiac arrest

Note: for analysis of clinical outcomes in HCM cases, HCM diagnosis was excluded from all of the above composite outcomes.

**EMC cohort methods**

**Clinical outcomes**

Three endpoints were predefined and evaluated in the Erasmus Medical Centre (EMC) cohort of 214 sarcomere-positive relatives of HCM probands.

1. HCM diagnosis
2. LV maximum wall thickness (maxLVWT) at last available transthoracic echocardiogram (TTE) or cardiac magnetic resonance imaging (CMR). For participants who underwent cardiac transplantation and/or septal reduction therapy, the last available imaging prior to cardiac transplantation and/or septal reduction therapy was used. Given the higher precision of CMR to assess wall thickness, maxLVWT from CMR was used whenever available unless CMR performed more than 5 years before the most recent TTE.
3. Time to first major adverse clinical event, a composite of myectomy therapy, alcohol ablation, cardiac transplantation, sustained ventricular arrhythmia, sudden cardiac death, or appropriate defibrillator shock.

**Statistical analysis**

The association between PGS_HCM_ and HCM was assessed using a Wald logistic mixed-effects model using the *glmm.wald()* function from R-package GMMAT (v.1.3.2)^4^, adjusting for fixed-effects of sex, age, age^2^ and PCs 1-4. To account for the between-sample relatedness, we incorporated a genetic relatedness matrix (GRM), estimated using GCTA (v.1.92.4 beta)^5^, as a random effect. The association of PGS_HCM_ with maxLVWT was assessed using a linear mixed-effects model implemented in function *lmekin()* from R-package coxme (v.2.2-17), adjusting for sex, age at imaging, age at imaging^2^, imaging modality (CMR vs TTE), ancestral PCs 1 to 4, and the GRM. The association of PGS_HCM_ with incident major clinical events was assessed using a Cox proportional hazards mixed-effects model using function *coxme()* from R-package coxme. Time 0 was set to birth in the Cox model and study participants were censored at the time of last clinical follow up. Sex, ancestral PCs 1 to 4 and the GRM were again included as covariates. *MYH7* rare variant genotype was more prevalent among samples with higher PGS_HCM_ (**Supplementary Table 1**); despite rare variant genotype not being found associated with any of the three endpoints, we performed sensitivity analyses adjusting for rare variant gene for each endpoint.

Primary analyses were performed using PGS_HCM_ as a continuous variable. In secondary analyses, we split PGS_HCM_ into quintiles and performed analyses testing top PGS quintile versus bottom PGS quintile, and top quintile versus median quintile. These analyses were performed to assess effect sizes between meaningful extremes of the PGS distribution.

**Variant pathogenicity**

***UK Biobank***

Pathogenic rare variants in 8 HCM-causing genes (*MYBPC3, MYH7, TNNT2, TNNI3, TPM1, ACTC1, MYL3,* and *MYL2*) were identified from whole exome sequencing data using the following approach^1^. Variants 100bp up or downstream of genes with a minor allele frequency of <0.1% in gnomAD and UKB were extracted. Splice region variants reported as pathogenic in ClinVar were manually curated for functional evidence of splicing^6^. LOFTEE^7^ was used to identify low confidence predicted loss of function (pLOF) variants, and all remaining pLOF variants were annotated for prediction of nonsense mediated decay (NMD) escape. The variants were then filtered for disease-causing mechanism and met a filtering allele frequency of <0.00004 in gnomAD: all MYBPC3 protein-altering variants were kept, and for the other 7 genes only variants influencing gene product structure or level were retained. Finally, the variant list was shorted to only include variants that would be called pathogenic or likely pathogenic in a patient with HCM using CardioClassifier^8^ and ClinVar, with manual curation of variants if they had evidence of pathogenicity.

***100K Genomes Project***

Pathogenic rare variants in 8 HCM-causing genes (*MYBPC3, MYH7, TNNT2, TNNI3, TPM1, ACTC1, MYL3,* and *MYL2*) were identified from whole genome sequencing data. Variants 100bp up or downstream of genes with a minor allele frequency of <0.1% in gnomAD and GeL were filtered for pathogenicity using ClinVar (annotated as pathogenic or likely pathogenic for HCM from multiple submitters without conflict) or CardioClassifier^8^, and for disease-causing mechanism (variants influencing gene product structure or level in *MYH7, TNNT2, TNNI3, TPM1, ACTC1, MYL3,* and *MYL2*).

***Royal Brompton Hospital***

HCM probands were sequenced using the Illumina Trusight Cardio Sequencing Kit on Illumina MiSeq and NextSeq platforms and a custom Agilent SureSelect cardiac gene panel on the Life Technologies SOLiD 5500xl platform. Rare variants were defined as having a mean allelic frequency of less than 1x10^-4^ in ExAC, and were required to be protein-altering (missense, nonsense, frameshift, in-frame indels, and essential splice site) and with high quality calling (PASS filter). Rare variant pathogenicity was determined following ACMG variant classification guidelines^9,10^ and incorporated the use of the CardioClassifier resource^8^.

***Erasmus Cohort***

Apart from genotyping using the Global Screening Array (in the research setting), all Erasmus cohort samples also underwent sequencing or targeted genotyping in the context of clinical genetics assessments. All samples included in the present study were found to be carriers of class 4 or class 5 variants in sarcomere genes (*MYBPC3*, *MYH7, TNNT2, TNNI3, TPM1, ACTC1, MYL3,* and *MYL2*). Variant pathogenicity was assessed centrally according to the American College of Medical Genetics and Genomics and the Association for Molecular Pathology (ACMG/AMP) guidelines^9^ using an adapted version of the CardioClassifier resource^8^, as described previously^11^. Homozygous carriers and those carrying multiple pathogenic or likely pathogenic variants were excluded in the present analysis."

**Supplementary References**

1.	de Marvao, A.*, et al.* Phenotypic Expression and Outcomes in Individuals With Rare Genetic Variants of Hypertrophic Cardiomyopathy. *J Am Coll Cardiol* **78**, 1097-1110 (2021).

2.	Elliott, P.M.*, et al.* 2014 ESC Guidelines on diagnosis and management of hypertrophic cardiomyopathy: the Task Force for the Diagnosis and Management of Hypertrophic Cardiomyopathy of the European Society of Cardiology (ESC). *Eur Heart J* **35**, 2733-2779 (2014).

3.	Ommen, S.R.*, et al.* 2020 AHA/ACC Guideline for the Diagnosis and Treatment of Patients With Hypertrophic Cardiomyopathy: Executive Summary: A Report of the American College of Cardiology/American Heart Association Joint Committee on Clinical Practice Guidelines. *Circulation* **142**, e533-e557 (2020).

4.	Chen, H.*, et al.* Control for Population Structure and Relatedness for Binary Traits in Genetic Association Studies via Logistic Mixed Models. *The American Journal of Human Genetics* **98**, 653-666 (2016).

5.	Yang, J., Lee, S.H., Goddard, M.E. & Visscher, P.M. GCTA: a tool for genome-wide complex trait analysis. *Am J Hum Genet* **88**, 76-82 (2011).

6.	Jaganathan, K.*, et al.* Predicting Splicing from Primary Sequence with Deep Learning. *Cell* **176**, 535-548.e524 (2019).

7.	Karczewski, K.J.*, et al.* The mutational constraint spectrum quantified from variation in 141,456 humans. *Nature* **581**, 434-443 (2020).

8.	Whiffin, N.*, et al.* CardioClassifier: disease- and gene-specific computational decision support for clinical genome interpretation. *Genetics in Medicine* **20**, 1246-1254 (2018).

9.	Richards, S.*, et al.* Standards and guidelines for the interpretation of sequence variants: a joint consensus recommendation of the American College of Medical Genetics and Genomics and the Association for Molecular Pathology. *Genet Med* **17**, 405-424 (2015).

10.	Walsh, R.*, et al.* Reassessment of Mendelian gene pathogenicity using 7,855 cardiomyopathy cases and 60,706 reference samples. *Genet Med* **19**, 192-203 (2017).

11.	Tadros, R.*, et al.* Shared genetic pathways contribute to risk of hypertrophic and dilated cardiomyopathies with opposite directions of effect. *Nat Genet* **53**, 128-134 (2021).
